# Albumin-dependent and independent mechanisms in the syndrome of kwashiorkor

**DOI:** 10.1101/2021.05.31.21257914

**Authors:** Gerard Bryan Gonzales, James M Njunge, Bonface M Gichuki, Bijun Wen, Moses Ngari, Isabel Potani, Johnstone Thitiri, Debby Laukens, Jill Vanmassenhove, Wieger Voskuijl, Robert Bandsma, James A Berkley

## Abstract

The syndrome of kwashiorkor is a striking phenotype of childhood severe malnutrition (SM) comprising oedema, fatty liver, and skin and hair changes. Despite high fatality, the aetiology and pathophysiology of kwashiorkor remain enigmatic, including the role of serum albumin on oedema development. Here, we demonstrate that serum albumin is associated with the presence and severity of oedema among severely malnourished children. Further, in two independent cohorts of children in Malawi and Kenya, we show albumin-independent mechanisms are associated with oedema in SM, including oxidative stress and extracellular matrix (ECM) remodelling. Plasma concentrations of ECM-related proteins: lumican, podoplanin, lymphatic vessel endothelial hyaluronan receptor 1 (LYVE1) and matrix metalloproteinase (MMP)2 were associated with kwashiorkor. We therefore conclude that the pathophysiology of kwashiorkor has both albumin-dependent and independent mechanisms. We discuss the ways in which albumin-independent mechanisms may explain the clinical features observed in kwashiorkor.

## Introduction

The syndrome of kwashiorkor (a.k.a oedematous severe malnutrition) is a striking phenotype of childhood malnutrition comprising oedema, fatty liver, hair depigmentation, a desquamating skin rash and behavioural changes^1^. It is distinct from the syndrome of marasmus (a.k.a severe wasting), characterised by low weight-for-height (<-3 z scored from 2006 WHO standards), low (<115 mm) mid-upper arm circumference (MUAC), visible atrophy and loose skin. Kwashiorkor and marasmus may co-occur (i.e. marasmic-kwashiorkor). Whilst both phenotypes are assumed to result from inadequate nutritional intake, the aetiology of the oedematous phenotype remains elusive^1^. Several hypotheses on the pathophysiology of kwashiorkor have been proposed but none are supported by robust epidemiological or mechanistic evidence ^1^.

The earliest explanation of kwashiorkor was a low protein diet leading to hypoalbuminemia causing pathognomonic oedema ^2^. However, in 2007, prospective assessment of food and nutrient intake in a population at risk for kwashiorkor in India and Malawi found no association between measured protein intake and kwashiorkor^3, 4^. Furthermore, oedema in SM is observed to resolve independently of protein intake^5^ and without an increase in serum albumin ^6^. However, the latter has been questioned upon reanalysis of the original data suggesting serum albumin could have increased in these earlier studies ^7^.

Circulating essential amino acid (EAA) concentrations, particularly sulphated AAs, are reported to be lower in kwashiorkor compared to marasmus ^8, 9^. Some of the clinical characteristics of kwashiorkor are similar to those occurring in methionine deficiency, notably the skin lesions and reduced plasma glutathione levels. Furthermore, supplementation study of cysteine has been associated with faster resolution of oedema compared to alanine supplementation ^10^. However, supplementation of methionine itself did not yield the same effect ^11^, nor did supplementation of an antioxidant mixture containing N-acetylcysteine prevent the development of kwashiorkor ^12^. Recently, DNA hypomethylation has been reported in kwashiorkor relative to marasmus during active disease, but not in subsequently recovered cases in adulthood, which the authors attribute to reduced concentrations and methyl-flux of methionine ^13^. However, due to the absence of data from non-malnourished controls, the results could also suggest relative hypermethylation in marasmus. Both DNA hyper and hypomethylation have been observed in caloric restriction and ageing ^14, 15^. The conclusion of hypomethylation in kwashiorkor, instead of hypermethylation in marasmus, was based on a prior observation that methyl-flux from methionine is reduced in kwashiorkor. Thus, the role of methionine deficiency in kwashiorkor remains unclear.

Oxidative stress has been suggested as an alternative hypothesis. The “free radical hypothesis” proposed increased production of free radicals accompanied by a reduction in anti-oxidative mechanisms, leading to oedema, skin and hair changes and fatty liver ^16^. Low anti-oxidative capacity was attributed to monotonous diets lacking several micronutrients and minerats (riboflavin, nicotinic acid, carotene, selenium, zinc) and sulphur AAs, needed for glutathione synthesis ^1, 16^. However, a trial of antioxidant supplementation containing riboflavin, vitamin E, selenium, and N-acetylcysteine for kwashiorkor prevention was ineffective ^12^; although, differences in oxidative stress were not measured in that trial. A pilot study of supplementation of glutathione and/or alpha-lipoic acid however improved survival and reduction of skin lesions in severe kwashiorkor ^17^. Exogenous toxicants, including aflatoxins from staple foods like maize often consumed in low and middle-incomes countries could also increase oxidative stress and explain the accumulation of liver fat in kwashiorkor. However, despite finding aflatoxins in biological samples of children with kwashiorkor ^18–22^, a causal link has not be demonstrated and study biases may have influenced results ^23^.

Endothelial dysfunction from disruption of sulphated glycosaminoglycans (GAGs) has also been proposed to cause kwashiorkor ^24^. However, congenital conditions linked with the inability to produce GAGs are not typically associated with oedema formation in affected children ^1, 25, 26^. Abnormal gut microbiota has also been implicated, although a recent analysis found no differences in the faecal metagenome between children with kwashiorkor and marasmus ^27^. A causal link between gut microbiome has been proposed from work in germ-free mice transplanted with microbiome from children with kwashiorkor, however oedema was not reported to have occurred in these animals. The study was not designed to distinguish whether effects relating to kwashiorkor from those of marasmus and so did not include faecal samples from children with marasmus ^28^. Children with SM in general have an altered gut metagenome composition compared to healthy children ^28, 29^, indicating that dysregulation of the microbiome may be a cause or consequence of SM, but this does not explain the pathophysiology of kwashiorkor.

Our aim was to determine albumin-independent differences in pathophysiology between the two SM phenotypes to inform targeted prevention and treatment strategies ^30, 31^.

## Results

### Low serum albumin is necessary but not sufficient to develop kwashiorkor

We initially determined the association between serum albumin concentration and kwashiorkor using data from a clinical trial of reformulated therapeutic milk among hospitalized children with severe acute malnutrition conducted in Malawi and Kenya (the discovery cohort) ^32^. Within this trial, 79% (662/843) of the children had admission serum albumin data with a median (interquartile range; IQR) of 34g/L (IQR 24 – 40). HIV, older age, and enrolment at the Malawi site were associated with lower albumin concentrations, while breastfeeding, pre-existing heart disease, and presenting with severe pneumonia or diarrhoea were associated with higher albumin (all p<0.05) in multivariable analysis. Serum albumin concentration was negatively associated with the presence of kwashiorkor (aOR = 0.75 [95% CI: 0.71, 0.78] per g/L, p < 0.001). We observed a significant decline in serum albumin concentration with increasing oedema severity graded according to the WHO classification (Figure 1a). Almost all children with kwashiorkor had serum albumin levels below 35g/L, but many children with similarly low levels of serum albumin did not have oedema or other features of kwashiorkor (Figure 1a,b). In a separate external validation cohort children with SM recruited in hospital after stabilization in another previous clinical trial in Kenya ^33^, serum albumin concentrations were significantly lower in children presenting with kwashiorkor compared to those with marasmus (Figure 1c) (aOR = 0.92 [95% CI: 0.87, 0.96], p = 0.001). Oedema grading data were not collected for this study.

**Figure 1.**
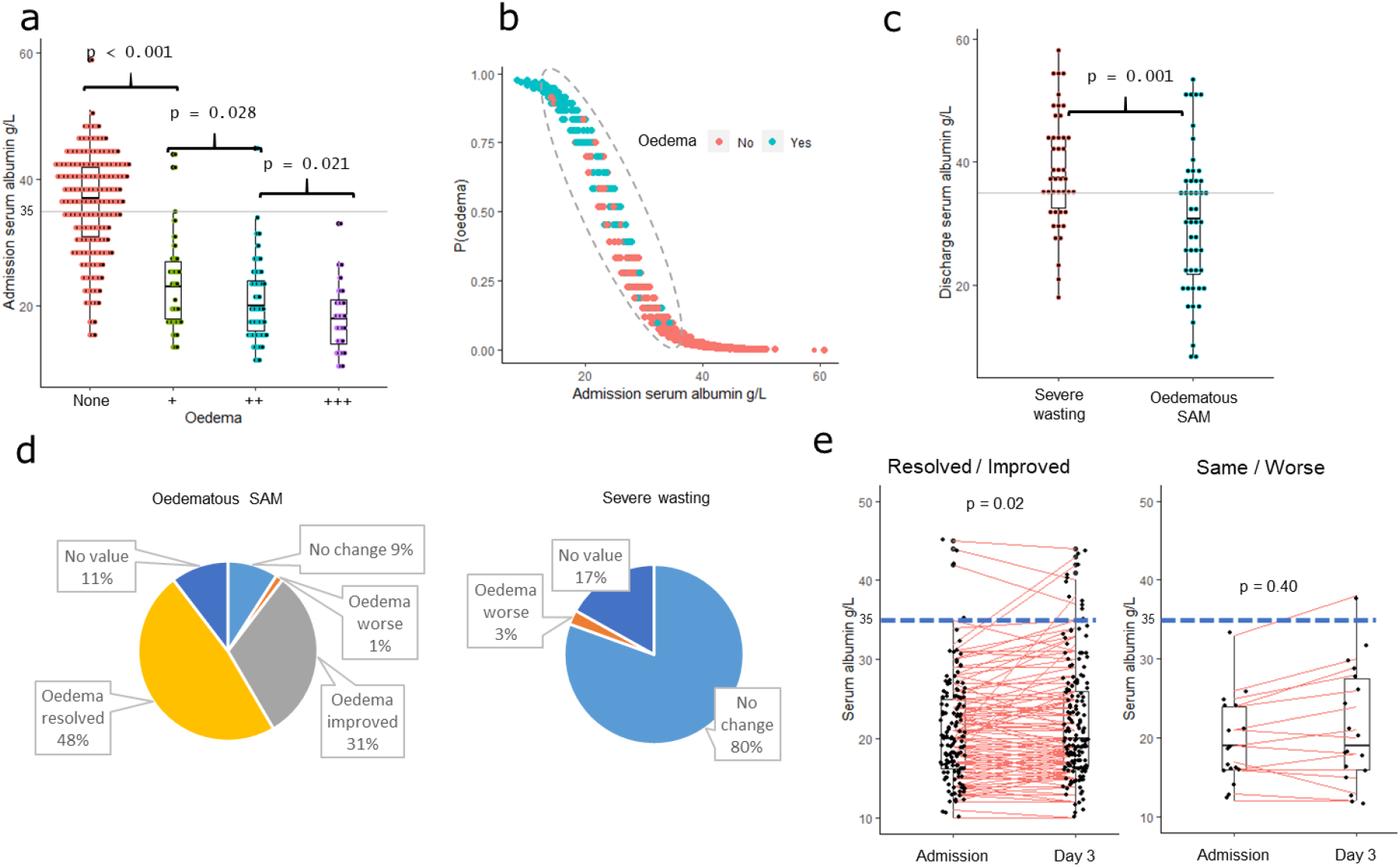
(a) Association between serum albumin concentration and degree of oedema severity in the F75 reformation clinical trial^32^: “None” means no oedema (marasmus) whereas “+” means oedema on both feet; “++” – oedema in both feet and legs; “+++” – oedema in both feet, legs, arms, hands and face. Oedema severity was assessed by trained clinicians following World Health Organization guidelines; (b) probability of presenting with oedema based on serum albumin concentration at hospital admission in the discovery cohort: green and red dots indicate those that presented with or without oedema respectively; (c) difference in serum albumin concentration between kwashiorkor and marasmus in the validation cohort; (d) distribution of oedema status after 3 days of hospitalization in the F75 reformulation clinical trial; (e) changes in serum albumin concentration among kwashiorkor during admission and after 3 days of hospitalization

Oedema resolved within 3 days of hospitalization in almost half (48%) of children admitted with kwashiorkor in the discovery cohort and oedema had improved in 31%. A small proportion (3%) of those admitted with marasmus developed oedema during treatment (Figure 1d). Among children whose nutritional oedema resolved or improved, there was a small increase in serum albumin concentration during 3 days of hospitalisation (0.68 g/L mean increase, p=0.02), whereas serum albumin remained unchanged among those whose oedema did not improve (Figure 1e). However, despite the small increase serum albumin among children whose oedema resolved or improved, concentrations at 3 days (median 20 g/L, IQR 16.5 – 26 g/L) were still far below clinically recognized norms in children (34 – 54 g/L). Adjusting for regression to the mean indicated no differences in changes in serum albumin between those without oedema and those with oedema which either improved (p = 0.93) or worsened (p = 0.38). These findings strongly suggest that although low serum albumin is associated with kwashiorkor, other factors play an essential role in the pathophysiology of the kwashiorkor phenotype.

### Selection of a sub-population matched on serum albumin levels

To determine factors associated with kwashiorkor in conjunction with low albumin, we further selected children from the F75 reformulation trial discovery cohort (Figure 2). This sub-cohort comprised children with kwashiorkor and marasmus who had been matched on exact serum albumin levels. In the discovery cohort, age and sex distributions were similar in oedematous and non-oedematous groups. Mid-upper arm circumference (MUAC) was higher among children with kwashiorkor (p < 0.001), whereas HIV was more prevalent among children with marasmus (p < 0.001). A greater proportion of oedematous children were recruited in Malawi than in Kenya.

**Figure 2.**
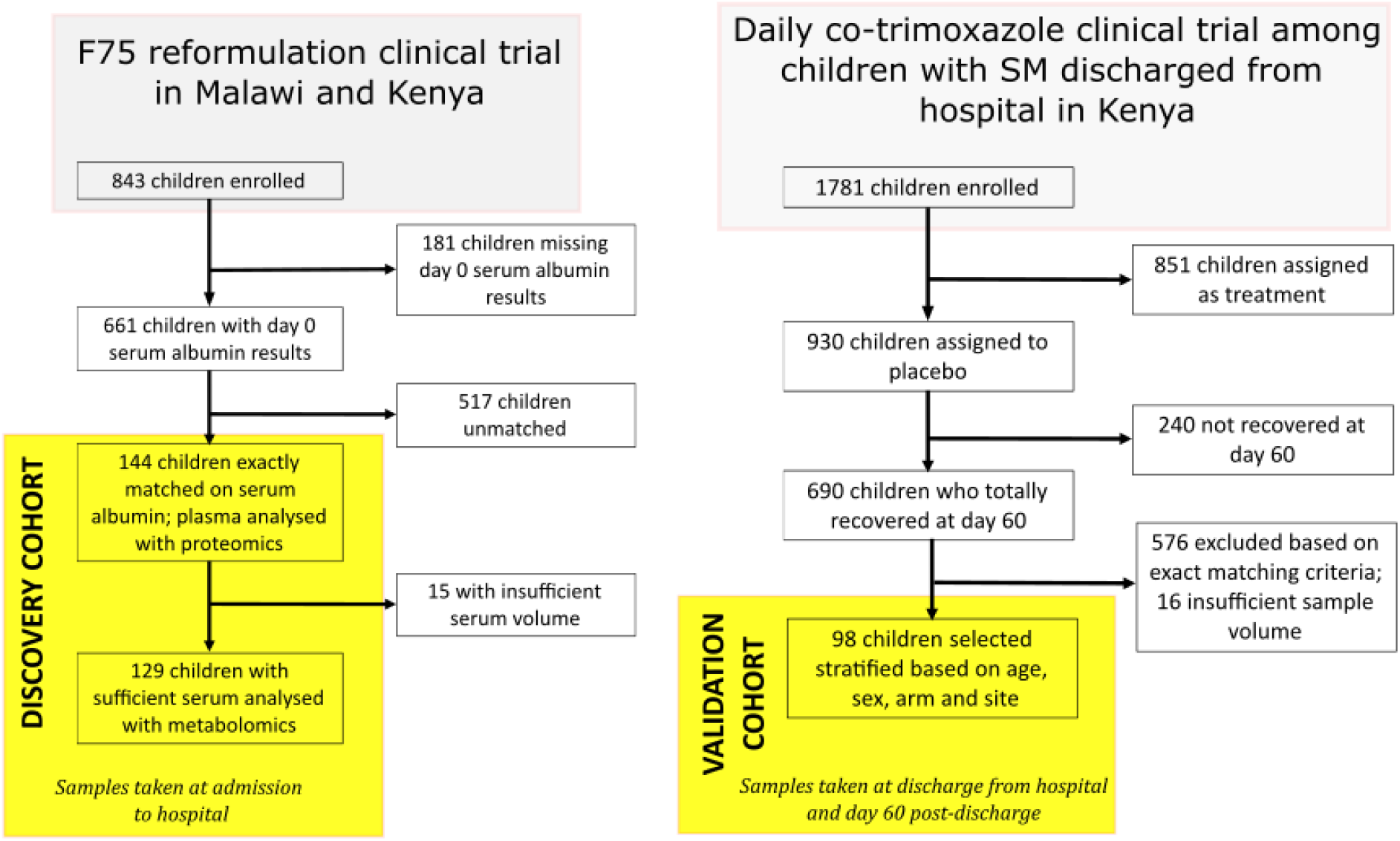
Recruitment flow diagram for the discovery and validation cohorts

To validate the results obtained from the discovery sub-cohort, a validation sub-cohort was selected from the validation set from the trial in Kenya, with kwashiorkor and marasmus matched on age, sex, site of recruitment and sex, but not matched for serum albumin concentration (more details in the Methods section) (Figure 2). In the validation cohort, 47 children with marasmus and 51 with kwashiorkor were selected based on 25 strata. As with the discovery cohort, serum albumin was higher in marasmus than in kwashiorkor thus, for succeeding analyses, variables were normalized by serum albumin concentration in order to assess differences occurring independently of serum albumin concentrations to harmonize approaches with the discovery cohort. The baseline description of both cohorts is presented in Table 1 and flowcharts of the selection for both cohorts are presented in Figure 2.

**Table 1.** Admission characteristics of the children used in this study from the two independent cohorts

|  | Discovery cohort |  |  | Validation cohort |  |  |
| --- | --- | --- | --- | --- | --- | --- |
| | Marasmus | Kwashiorkor | $p^*$ | Marasmus | Kwashiorkor | $p^*$ |
| $N$ | 72 | 72 | / | 47 | 51 | / |
| Median age (mo.)<br>at admission [IQR] | 18<br>[12 – 27] | 24<br>[15 – 31] | 0.92 | 16<br>[13 – 24] | 20<br>[17 – 25] | 0.08 |
| Girls $n$ (%) | 34 (47) | 36 (50) | 0.73 | 18 (38) | 21 (41) | 0.83 |
| MUAC, cm mean ± SD<sup>@</sup> | 10.3 ± 1.4 | 11.8 ± 1.7 | <0.001 | 10.8 ± 0.5 | 12.6 ± 0.9 | <0.001 |
| HIV status $n$ (%) | | | <0.001 | | | / |
| Positive | 36 (50) | 14 (19) |  |  |  |  |
| Negative | 32 (44) | 54 (75) |  | 47 (100) | 51 (100) |  |
| Unknown | 4 (6) | 4 (6) |  |  |  |  |
| Weight-for-length<br>z-score ± SD | -4.1 ± 1.5 | / | / | / | / | / |
| Recruitment<br>country $n$ (%) | | | <0.001 | | | 1 |
| Kenya |  |  |  |  |  |  |
| Coast Provincial<br>General Hospital | 26 | 6 |  | 26 | 29 |  |
| Kilifi County<br>Hospital | 22 | 22 |  | 4 | 5 |  |
| Malindi sub-<br>county Hospital |  |  |  | 5 | 5 |  |
| Mbagathi Hospital<br>Nairobi |  |  |  | 12 | 12 |  |
| Malawi |  |  |  |  |  |  |
| Queen Elizabeth<br>Central Hospital | 24 | 44 |  |  |  |  |
| Mean serum<br>albumin (g/L) ± SD | 24.4 ± 4.5 | 24.4 ± 4.5 | 1 | 38.6 ± 0.9 | 30.3 ± 1.2 | <0.001 |

\*Marasmus vs Kwashiorkor per cohort, <sup>@</sup>Mid-upper arm circumference

### Plasma lipids and extracellular matrix proteins are albumin-independent factors associated with kwashiorkor

To discover albumin-independent protein and metabolite factors associated with kwashiorkor, plasma samples from the discovery sub-cohort were subjected to untargeted liquid chromatography tandem mass tag (TMT)-based proteomics and targeted metabolomics (Biocrates™ p 180). After data cleaning and pre-processing, 187 out of 456 proteins and 155 out of 205 metabolites were retained for further analysis. A complete list of annotated proteins and targeted metabolites and their association with nutritional oedema is provided in Supplementary table S1. Nine metabolites including 1 phosphatidylcholine (PC), 6 lysoPC, and 1 sphingomyelin species, apolipoprotein C-I (Apo C1) and 3 extracellular matrix protein (ECM)-related proteins (lumican, inter-alpha-trypsin inhibitor heavy chain H2 [ITIH2] and histidine-rich glycoprotein [HRG]) were positively associated with kwashiorkor compared to marasmus cases matched by exact serum albumin levels (Figure 3a). Furthermore, all these metabolites and proteins were associated with increasing severity of oedema (Figure 3b).

**Figure 3.**
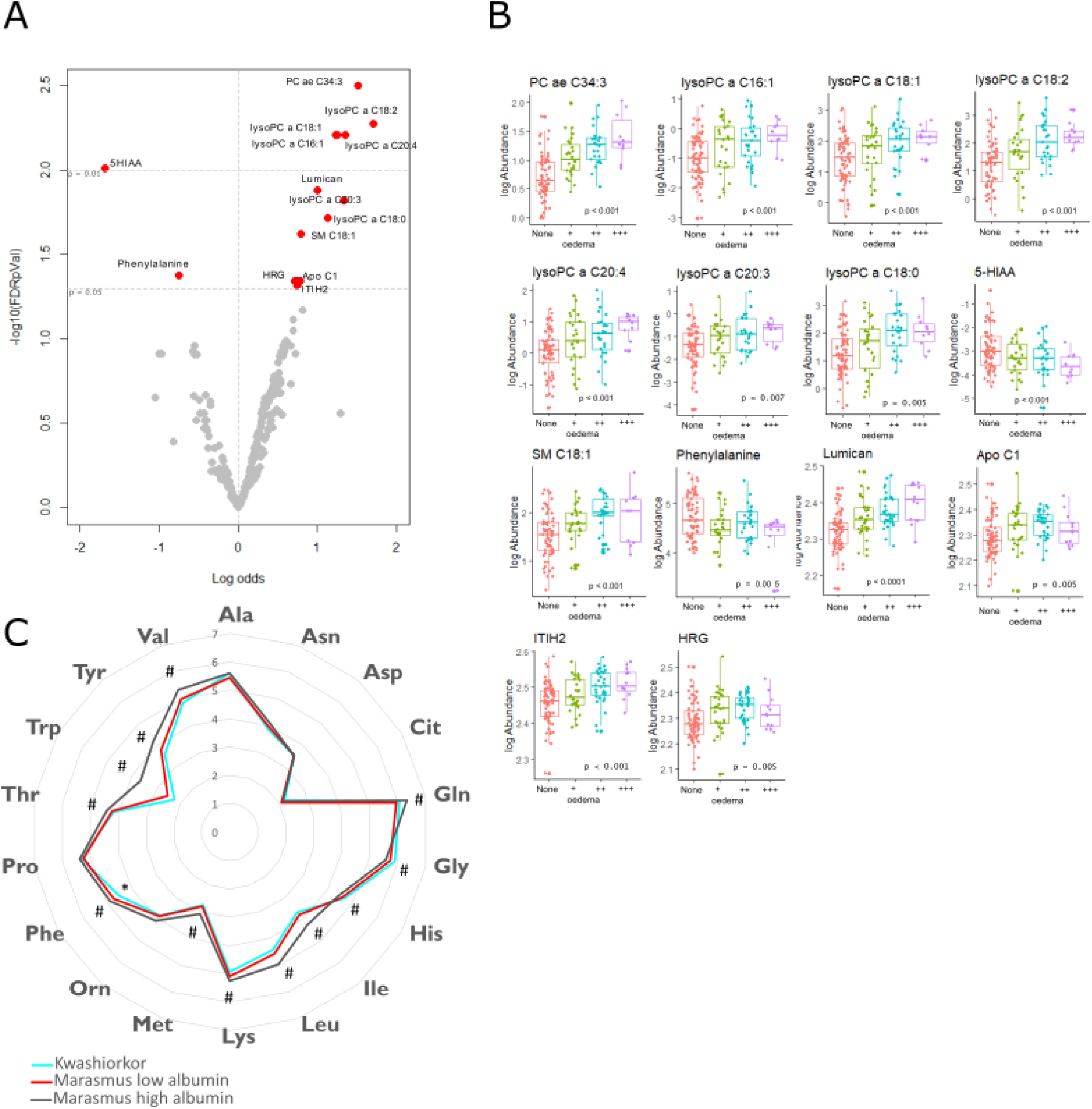
Differential abundance of proteins and metabolites between kwashiorkor and marasmus. (A) Volcano plot showing the log odds ratio (x-axis) and –log p value after false-discovery rate adjustment of plasma proteins (y-axis). False discovery adjustment was performed separately for proteins and metabolites. The horizontal line signify the FDR p = 0.05 and p = 0.01 cut-offs, whereas vertical broken line signify log odds ratio = 0. Features on the upper right quadrant represent those with FDR-corrected p values < 0.05, and are associated with kwashiorkor compared to marasmus phenotype, whereas those in the upper left quadrant are associated with marasmus. Estimates were obtained using conditional logistic regression adjusting for age, sex, HIV status and site of recruitment stratified for admission serum albumin concentration. (B) Boxplot showing the association between the plasma concentrations of significantly associated features and the degree of oedema severity. “None” means no oedema (marasmus) whereas “+” means oedema on both feet; “++” – oedema in both feet and legs; “+++” – oedema in both feet, legs, arms, hands and face. Oedema severity was assessed by trained clinicians following World Health Organization guidelines; p values were estimated using ordinal logistic regression adjusted for age, sex, HIV status, site of recruitment and serum albumin concentration. (C) Radar plot comparing plasma amino acid content in kwashiorkor and marasmus both with matched serum albumin and serum albumin > 35g/L (high albumin). * denotes difference at p <0.05 between kwashiorkor and marasmus matched with serum albumin. # denoted different at p < 0.05 between kwashiorkor and marasmus with serum albumin >35g/L (high albumin)

### Plasma phenylalanine and 5-hydroxyindoleacetic acid (5-HIAA), but not methionine, distinguish SM phenotype

Plasma levels of phenylalanine (aOR = 0.47 [95% confidence interval: 0.29, 0.77]) and 5-HIAA (aOR = 0.18 [95% confidence interval: 0.07, 0.47]) were negatively associated with kwashiorkor. On the contrary, methionine levels were not associated with SM phenotype (aOR = 0.67 [95% confidence interval: 0.42, 1.08]) nor with oedema severity. In fact, an almost complete overlap between each free amino acid concentration was observed between kwashiorkor and marasmus (Figure 3c). Although 5-HIAA was negatively associated with kwashiorkor, plasma levels of the other members of the tryptophan pathway, i.e. tryptophan, serotonin and kynurenine, were not (Supplementary figure S1). Comparing kwashiorkor with children with marasmus but with serum albumin > 35g/L (high albumin), almost all measured amino acid concentrations were higher (p < 0.05) in the high albumin-marasmus group compared to kwashiorkor, indicating an association between serum albumin concentration on measured plasma amino acid content. Asparagine, aspartic acid, citrulline, ornithine and proline concentrations were not different between kwashiorkor and marasmus, whereas glycine and histidine were significantly lower in the high albumin-marasmus group compared to kwashiorkor.

### Multi-omics co-expression network analysis uncovers albumin-independent mechanisms associated with oedema in SM

Co-expression network analysis of integrated proteomics and metabolomics data identified fifteen modules of strongly correlated features (Figure 4a). Features not clustering with any module are presented in module ME7. Module membership is presented in Supplementary Table S2. A general description of the membership of each module is shown in Figure 4a. Briefly, PC modules (ME1, 3, 4, 9, 10, 13 and 14) were clustered based on the lengths of their side chains, bonding to the glycerol backbone (either ether or ester-linked) and the degree of unsaturation (average number of double bonds). Most amino acids, lysoPCs and sphingomyelines clustered in separate modules (ME5, ME11 and ME12, respectively). ME6 contained unsaturated acylcarnitines, whereas saturated acylcarnitines were placed with other unassigned metabolites in ME7. Protein-rich modules were characterized by gene-ontology (GO) enrichment analysis ^34^. M8 was composed of proteins associated with oxidative stress whereas M16 with immunoglobulins. The two large protein modules were ME2 and M15. Although closely related, ME2 was mainly composed of proteins associated with immune response and complement activation whereas ME15 was mainly composed of proteins associated with lipid transport and metabolism, and extracellular matrix remodelling.

**Figure 4.**
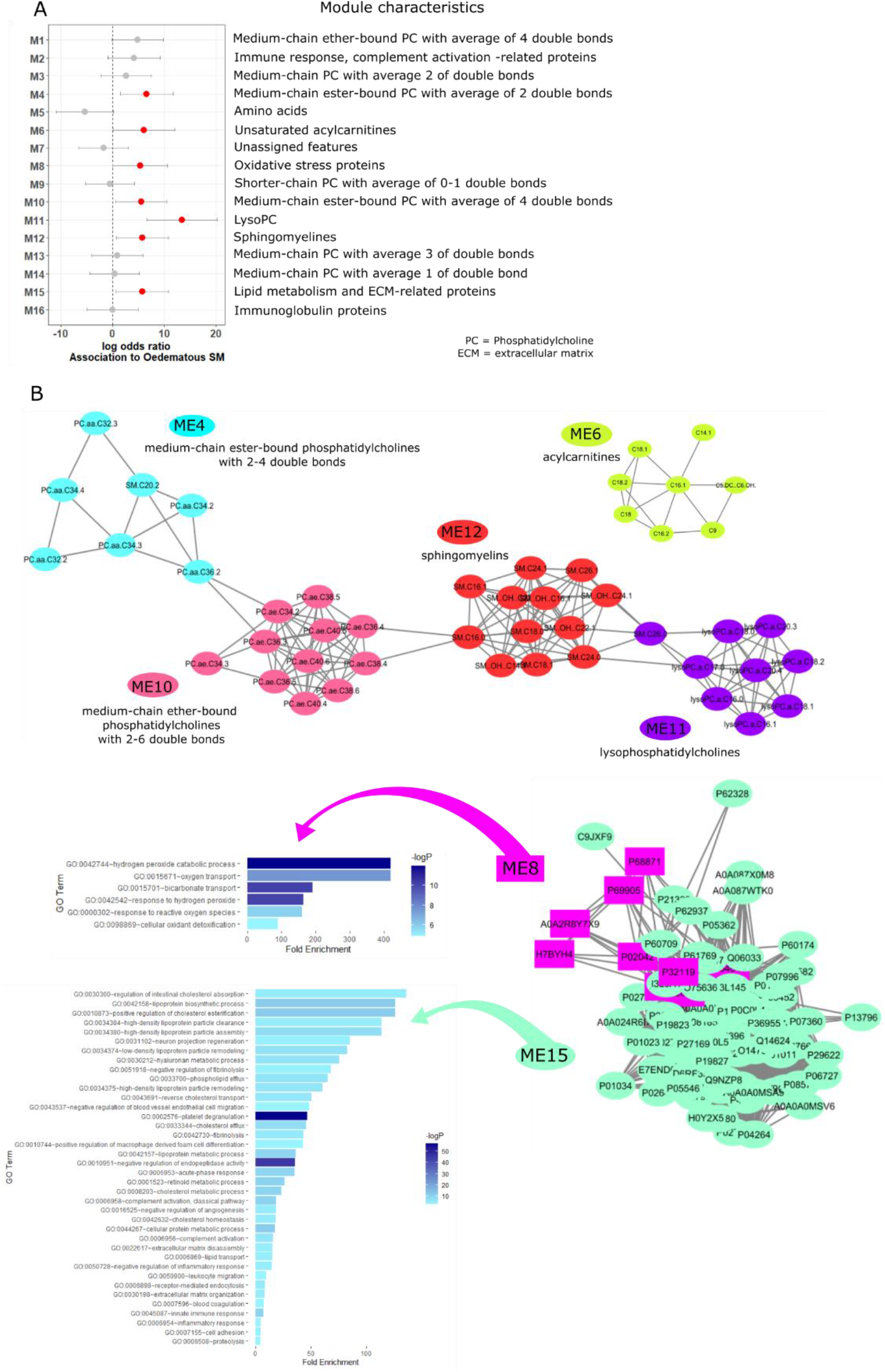
Multi-omics weighted co-expression network analysis. (A) Plot showing log odds ratio with binary outcome (kwashiorkor vs marasmus) as response variable and each module’s eigenvector as independent variable adjusted for age, sex, HIV status, site of recruitment and stratified based on admission serum albumin using conditional logistic regression. Side bars indicate 95% confidence interval. Red dots denote modules significantly (p < 0.05) associated with kwashiorkor. (B) Network depiction of the modules associated with kwashiorkor showing the members of each module and their module characteristics. Network structure was obtained using the WGCNA package in R and visualized using Cytoscape version 3.8.2.

Each module is characterised by an eigenvector (E^(q)^, which is the 1^st^ principal component of module q that represents the overall behaviour of the module^35^. Regressing E^(q)^ to a binary outcome (kwashiorkor or marasmus) adjusting for age, sex, HIV and recruitment site, stratified by each value of admission serum albumin, we found seven modules (ME4, 6, 8, 10, 11 and 15) to be significantly associated with kwashiorkor (Figure 4a,b).

Most of the associated modules were composed of lipids (ME4, 6, 10-12). However, there was a preferential increase in plasma concentration of unsaturated ester-bound PCs (ME4, 10) compared to ether-bound PCs (ME1) and PCs with lower degree of unsaturation (ME9) in kwashiorkor. Modules containing both ester- and ether-bound PCs were also not associated with kwashiorkor, further emphasizing the preference for esterified PCs. Plasma levels of unsaturated acylcarnitines (ME6) were also preferentially increased in kwashiorkor compared to saturated acylcarnitines (clustered in ME7). LysoPCs (ME11) and sphingomyelins (ME12) were found to be positively associated with kwashiorkor compared to marasmus. These positive associations of lipid modules to kwashiorkor are corroborated by the results of lipid specific associations (Figure 3a) and the positive association of proteins involved in lipid transport and metabolism (ME15). Proteins associated with oxidative stress (ME8) was also positively associated with kwashiorkor.

We further observed that proteins linked with extracellular matrix remodelling (ME15) were positively associated with kwashiorkor. This also corroborates with the results of protein specific associations (Figure 3a), showing that ECM-associated proteins lumican, ITIH2 and HRG were positively associated with kwashiorkor.

### Plasma markers of endothelial glycocalyx integrity are associated with oedema severity but not specifically with kwashiorkor

We quantified lumican using ELISA to validate the untargeted proteomics results in both discovery and validation cohorts. Lumican was found to be positively associated with kwashiorkor compared to marasmus (aOR = 1.49 [95% CI: 1.23, 1.79]) per µg/mL, adjusting for age, sex, HIV status and site of recruitment in agreement with the untargeted proteomics results (Figure 5a). Also consistent with untargeted proteomics findings, plasma lumican concentration was positively associated with increasing degree of oedema severity (p < 0.001) (Figure 5b). The positive association of plasma lumican and kwashiorkor was replicated in the validation cohort for lumican levels normalized on serum albumin concentration (p = 0.002) (Figure 5c). Comparing circulating lumican levels between discharge (still SM but free of underlying infections) and 60 days post-discharge (fully recovered from SM and infections) in the validation cohort, lumican was found to significantly increase (p < 0.001) among children with marasmus, whereas it decreased among children with kwashiorkor (p < 0.03).

**Figure 5.**
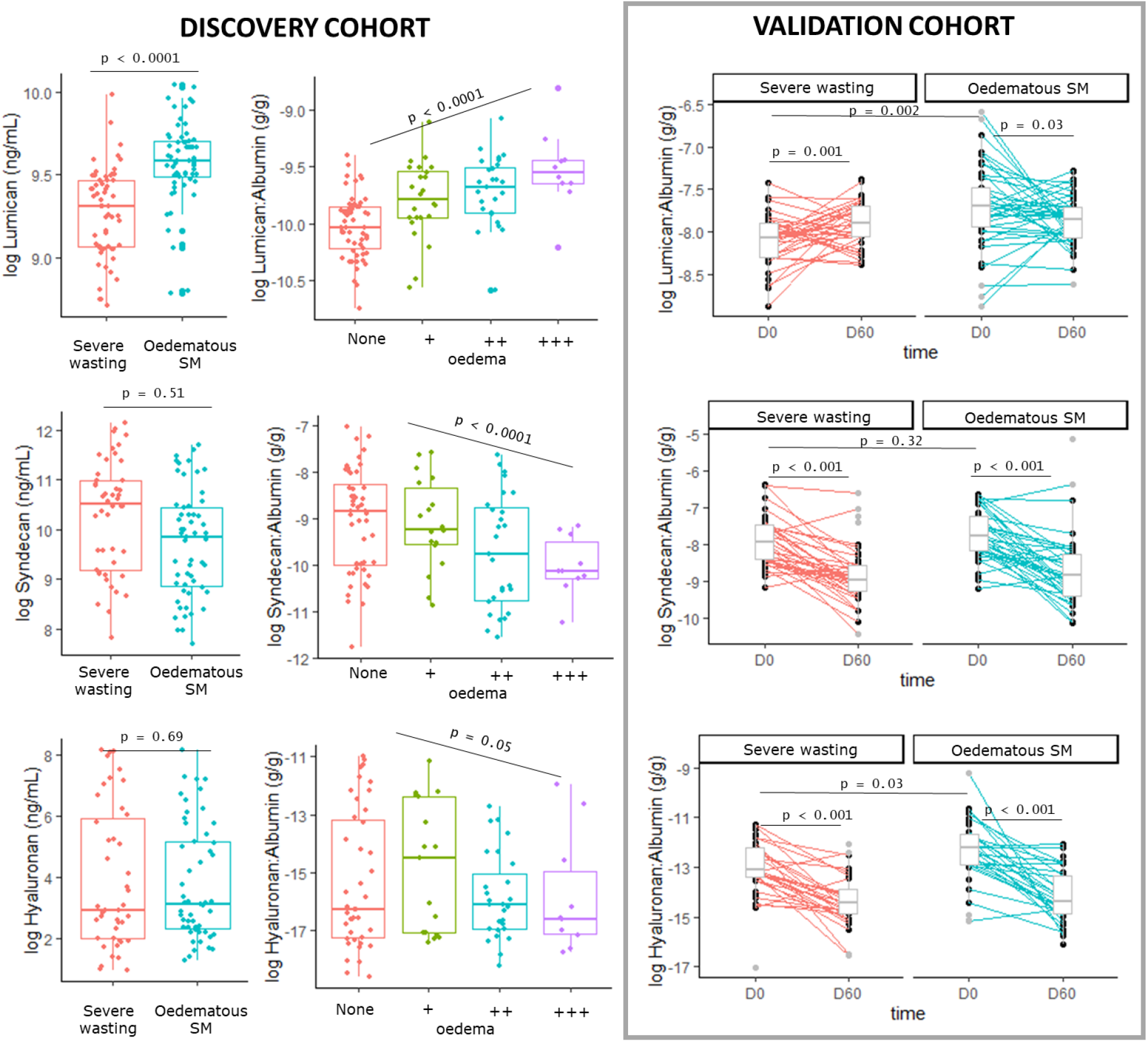
Association between SM phenotype and lumican and endothelial glycocalyx markers.

As lumican has been reported to adsorb on the endothelial glycocalyx ^36^, we reasoned that the increase in plasma lumican likely reflected a disruption of the endothelial glycocalyx. Hence, we determined the association between kwashiorkor and plasma endothelial glycocalyx (EG) markers. Of the several EG integrity markers reported in the literature, and given limited sample volumes, we prioritised analysis of two abundant markers in plasma previously reported to be increased in diseases with known EG dysfunction ^37^: syndecan-1 (Syn1), a proteoglycan bearing sulphated glycosaminoglycans (GAG), and hyaluronan (HA), a non-sulphated GAG.

Based on literature suggesting increased serum levels of EG markers in diseases associated with leakage of intra-vascular fluid and proteins leading to oedema, such as dengue^37^, we initially hypothesized that increased plasma levels of Syn1 and HA would be associated with kwashiorkor. Surprisingly, neither Syn1 nor HA were associated with kwashiorkor compared to marasmus in the discovery cohort (Figure 5d-i). Their plasma levels were however negatively associated with increasing degree of oedema severity. These results were replicated in the validation cohort, except for a modest but significant increase in HA among children with kwashiorkor (Figure 5i). Plasma levels of Syn1 and HA significantly decreased after 60 days post discharge among children with either initial phenotype who fully recovered from malnutrition without further acute illness following discharge.

### ECM remodelling is an albumin-independent mechanism in kwashiorkor

Plasma levels of matrix metalloproteinase (MMP)2 in the discovery cohort had a positive association with kwashiorkor (aOR = 1.89 [95% confidence interval: 1.38, 2.58]) per µg/mL, and was associated with severity of oedema (p < 0.0001) (Figure 6a). Because of this, we measured plasma levels of ECM remodelling regulators, i.e. MMP2, tissue inhibitors of MMP (TIMP)1, and TIMP2, and other ECM proteins (podoplanin and LYVE1) in the validation cohort, both at hospital discharge and at day 60 post-discharge among children who achieved full nutritional recovery.

**Figure 6.**
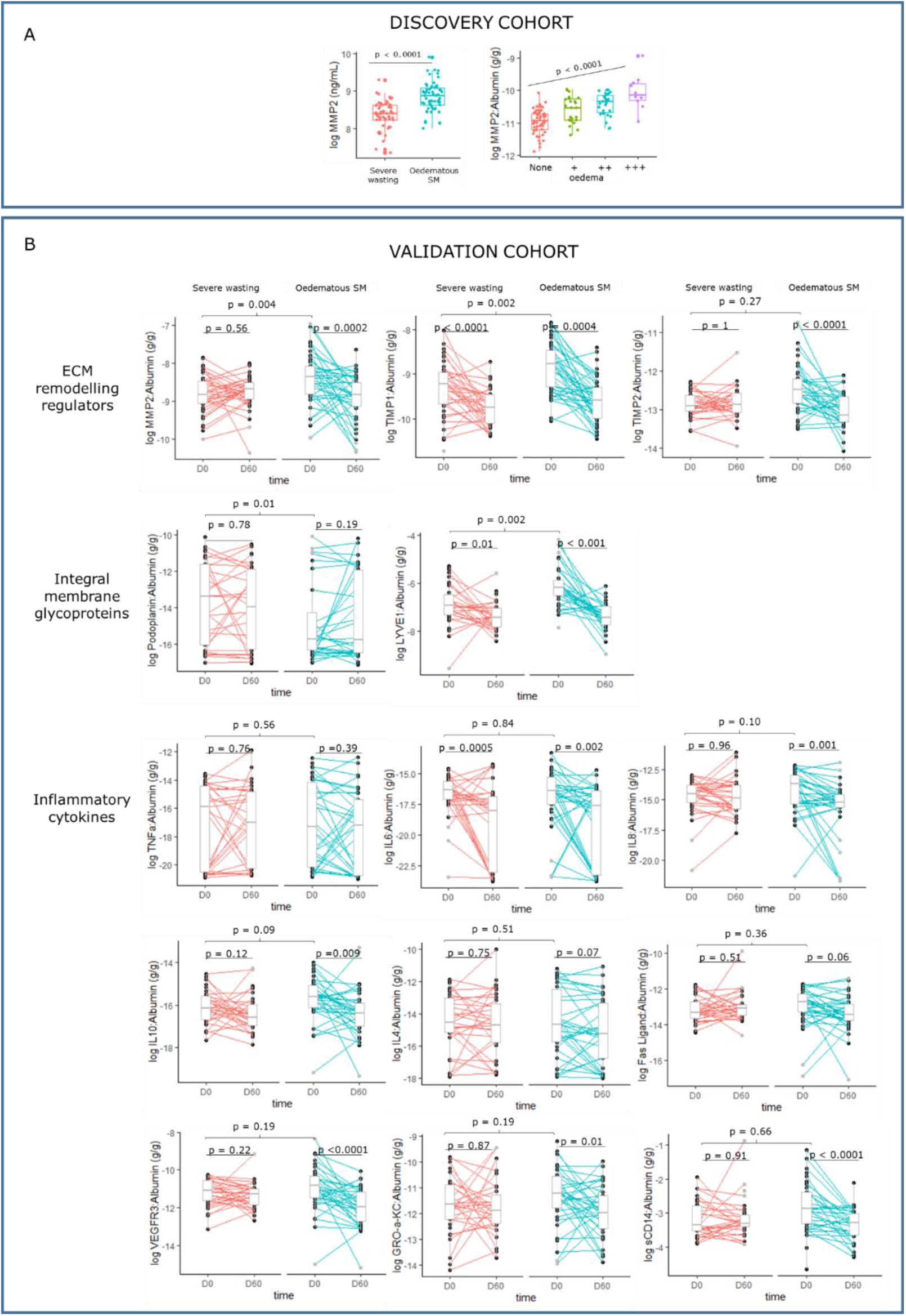
Association between SM phenotype and markers of ECM remodelling markers and systemic inflammatory.

ECM remodelling regulators MMP2 and TIMP1 were positively associated with kwashiorkor. Furthermore, plasma levels of these proteins significantly reduced during nutritional rehabilitation (Figure 6b). Apart from lumican, other ECM proteins were altered in plasma of children with kwashiorkor. Podoplanin, a mucin-like glycoprotein found in the alveoli, heart and lymphatic vascular system, was negatively associated, whereas lymphatic vessel endothelial HA receptor 1 (LYVE1) was positively associated with kwashiorkor. Plasma levels of LYVE1 also reduced during nutritional rehabilitation in both the kwashiorkor and marasmus phenotypes, but podoplanin remained unchanged.

As ECM remodelling is activated by inflammation, we further measured key plasma markers of systemic inflammation in both SM phenotypes. None of the inflammatory cytokines measured were differentially abundant between kwashiorkor and marasmus cases matched for serum albumin, suggesting inflammatory response-independent mechanisms were driving ECM remodelling in kwashiorkor. It can be observed however that plasma levels of IL8, IL10, VEGFR3, GRO alpha KC and sCD14 decreased significantly in plasma of validation cohort children with kwashiorkor during the 60 days post-hospital discharge but remained unchanged in marasmus. Plasma IL6 significantly decreased in both kwashiorkor and marasmus upon full nutritional and clinical recovery (Figure 6B).

## Discussion

There is a universal understanding within the medical literature of the role of serum albumin concentration on oedema formation in general, as explained by Starling forces. However, in hypoalbuminemic states such as the nephrotic syndrome and inflammation, evidence suggests that other factors beyond a decline in colloid osmotic pressure are also responsible for the altered fluid distribution^38^. The role that albumin plays in kwashiorkor has been a topic of debate among scientists, with some postulating a causal role of hypoalbuminaemia in its aetiology^7, 39^ while others rejecting their association ^6, 40^. In this study, we showed that kwashiorkor is associated with both albumin-dependent and independent mechanisms. We demonstrated for the first time a role of ECM degradation as an albumin-independent mechanism in the pathophysiology of kwashiorkor.

Our data from two independent cohorts show that serum albumin is lower among children with kwashiorkor compared to marasmus and negatively associated with the degree of oedema severity. In fact, there were almost no cases of kwashiorkor where serum albumin concentrations were more than 35 g/L in the discovery cohort. In the validation cohort who were enrolled towards the end of their hospital admission having initiated therapeutic feeding and no longer suffering acute infection, 35% of kwashiorkor cases had serum albumin levels above 35 g/L compared to 67% for the marasmus cases. These data provide the first physiological validation of the current WHO nutritional oedema grading system ^41^. However, in the absence of active acute infection (validation cohort), serum albumin concentrations overlapped more between groups compared to at admission to the hospital (F75 reformulation trial). Furthermore, we observed that resolution or improvement of oedema was accompanied by a small but not likely clinically relevant increase in serum albumin concentrations, agreeing with previous observations ^5, 6^. These led us to postulate that other factors beyond albumin play important roles in oedema formation in SM.

Using a targeted metabolomics approach, we previously showed that serum concentrations of 141 metabolites (including AAs) tended to be lower in kwashiorkor compared to marasmus in general, and not only AA ^42^. In this current study, we found that plasma AA profile was very similar between kwashiorkor and marasmus when matched for serum albumin concentrations, contrary to previous reports in which serum albumin concentrations were not considered ^8, 9, 42^. These data indicate an association between serum albumin and serum free AA concentrations. One explanation for this is water displacement by albumin. In conditions of low albumin content, water replaces the space that would have been occupied by albumin per volume of serum, thereby causing a dilution of polar metabolites. Hence, an apparently lower content per unit volume of polar metabolites is observed in kwashiorkor compared to marasmus with high serum albumin concentration. Another potential explanation is that the lower serum albumin concentration is a reflection of low concentrations of AA precursors needed for albumin synthesis. We therefore addressed these issues by matching for serum albumin when comparing kwashiorkor and marasmus in the discovery cohort and normalising for serum albumin in the validation cohort.

Our results also demonstrate that kwashiorkor is associated with increased levels of oxidative stress (increased hydrogen peroxide catabolism, oxygen transport, response to reactive oxygen species and cellular oxidant detoxification). However, whether increased oxidative stress causes or is a consequence of kwashiorkor requires further investigation. Nonetheless, we showed in this study that oxidative stress was increased in kwashiorkor independently of the plasma AA profile, which agrees with one of the postulates of the free radical hypothesis ^16^. However, we did not find evidence for an association between plasma levels of endothelial glycocalyx components, Syn1 and HA, and kwashiorkor. Instead we found other ECM proteins that are associated with kwashiorkor, such as lumican, LYVE1, podoplanin and MMP2. It is important to note however that endothelial glycocalyx components are also structurally linked to ECM proteins.

Lumican is a leucine-rich proteoglycan with keratan sulphate, a sulphated glycosaminoglycan (sGAG), side chains and is a major component of corneal, dermal and muscle connective tissue. Downregulation of lumican results in skin fragility and laxity, and corneal opacity ^43^. Hence, degradation of lumican could explain the skin changes^44, 45^ and corneal opacity ^46, 47^ reported to occur in some children with kwashiorkor. Although, no study has linked lumican specifically with the type of skin changes in kwashiorkor, i.e. “flaky paint” or “peeling paint” dermatosis, reduction of lumican levels have been reported in skin diseases such as actinic keratosis and Bowen disease ^48^.

The lymphatic system could also be involved in the clinical manifestations of kwashiorkor, especially oedema. LYVE1 and podoplanin are both markers of lymphatic endothelial integrity predominantly expressed in lymphatic vessels ^49–51^. LYVE1 and podoplanin are essential for lymphatic system development and ablation of podoplanin and LYVE1 in transgenic mice resulted in diminished lymphatic transport and lymphedema ^52, 53^. Hence, differential plasma levels of these markers indicate that lymphatic system could be compromised in kwashiorkor, leading to poor fluid homeostasis. Degradation of ECM in the lymph may also explain the disturbances in lipid metabolism observed in kwashiorkor, being the main route for chylomicrons from enterocytes to reach the bloodstream ^54^. Interestingly, reduction of podoplanin by knock-out of T-synthase in mice caused fatty liver by chylomicrons being diverted directly to the portal vein instead of being transported to the bloodstream via the lymphatic system ^55^. It is however worth noting that liver steatosis is not a common observation among children with congenital intestinal lymphangiectasia, a disease associated with lymphatic damage^56^. Hepatic lipid accumulation in animal models of SM has so far been proposed to be related to mitochondrial dysfunction, peroxisomal damage^57, 58^ or choline deficiency ^59^. However, in these animal studies, rodents were fed low-protein diets which did not induce oedema. ECM degradation, especially in the lymph, could be a contributing mechanism to liver fat accumulation in kwashiorkor, which is testable. Analysis of post-mortem portal vein triglyceride content using minimally invasive tissue sampling strategies could test this hypothesis. Alternatively, high content of triglycerides, apolipoprotein B48 and lymphocytes in ascitic fluids could further demonstrate the role of lymphatic degradation in kwashiorkor. However, ascites in kwashiorkor may already indicate a more advanced condition or an entirely different clinical picture, which could influence this observation.

Both LYVE1 and podoplanin (via colocalization with CD44) are major HA receptors ^50, 60^. Although we did not find significant association between kwashiorkor and HA in the discovery cohort, plasma albumin-normalized HA was higher in kwashiorkor in the validation cohort, indicating higher HA shedding. Observed differences between our cohorts could be because children in the discovery cohort were both severely undernourished and clinically ill at the time of sampling, whereas children in the validation cohort were still severely undernourished but severe infection had resolved, micronutrients, protein and energy had been initiated, and they were tolerating therapeutic feeds, which is regarded as a sign of nutritional stabilisation.

Increased levels of plasma ECM were accompanied by increased plasma concentrations of MMP2, which is an active regulator of ECM remodelling. MMP2 is a 72 kDa type IV collagenase that is distributed in many tissues and associated with several serious diseases. Along with MMP9, it is also expressed in lymphatic endothelial cells ^61^ and plays a key role in lymphatic vessel formation ^62^. Oxidative stress is a strong activator of MMP2 ^63^, and reactive oxygen–nitrogen species-induced MMP2 activation has been shown to play major roles in various diseases such as cardiac injury ^64^, hepatic fibrosis ^65^ and atherosclerosis ^66^. Our findings therefore provide a plausible mechanism for previous data suggesting generalized loss of sGAG in kwashiorkor ^24, 67–70^. Our results suggest an MMP-induced degradation of the core ECM proteins that these sGAG are structurally linked to. We also observed that this disruption is not exclusive to sGAG-binding ECM but also to ECM proteins linked to non-sulphated GAGs, such as LYVE1 and podoplanin.

We did not find evidence of greater systemic inflammation in kwashiorkor compared to marasmus, when albumin was matched. This is surprising considering the well-described interaction among oxidative stress, inflammation and ECM remodelling ^71^. It is thus plausible that children with kwashiorkor have a predisposition to a heightened ECM remodelling given the same inflammatory insult experienced by children with marasmus. Hence, examining variability in ECM remodelling-associated genes, such as MMPs, would be a promising next step to understand the aetiology of kwashiorkor. These results also highlight the role of non-nutritional factors such as (epi)genetic factors in the aetiology of kwashiorkor.

Although our results do not give a definite answer to what causes kwashiorkor, we reveal important findings on the pathophysiology of oedema, especially the role that albumin plays. This finding is not only relevant to SM but also to other oedematous diseases such as nephrotic syndrome and sepsis. We recognize that matching patients by admission serum albumin to specifically examine albumin-independent mechanisms associated with kwashiorkor precludes comparison with the full spectrum of marasmus cases. Nonetheless, our results remained consistent in the validation cohort where we normalized by albumin rather than individually matched. A limitation of the study is the lack of data on the renal function, which has also been reported to be associated with LYVE1 ^72^, podoplanin ^73^ and lumican ^74^. Although it is plausible that any renal dysfunction in kwashiorkor could be linked with the degradation of renal ECM proteins.

Given these findings, we propose that the mechanism for oedema in kwashiorkor involves both reduced serum albumin concentration and increased oxidative stress leading to heightened ECM degradation resulting in (1) reduction in interstitial integrity and (2) impaired lymphatic integrity causing poor interstitial fluid drainage. The ECM is a highly dynamic structure in which components are continuously synthesized, degraded and regenerated ^75^. This potentially explains the resolution of oedema despite minimal increase in serum albumin concentrations. Hence, targeting restoration of ECM during treatment could help rapidly restore interstitial rigidity and lymphatic integrity allowing the drainage of interstitially displaced fluids back to circulation.

## Methods

### Overall study design

This study comprised of two separate nested case-control studies formed from a sub-selection of children with either kwashiorkor or marasmus from two clinical trials in Malawi and Kenya, as further described below. A hypothesis generating discovery cohort was used to explore albumin-independent mechanisms, which were then validated using the second cohort.

### Study population and setting

#### Discovery cohort

The discovery cohort was nested within a randomised controlled trial (NCT02246296) that aimed at determining the effect of a lactose-free, low-carbohydrate F75 milk formulated to limit carbohydrate malabsorption, diarrhoea and refeeding syndrome among children hospitalized for complicated SM in Queen Elizabeth Central Hospital in Malawi, and Kilifi County Hospital and Coast General Teaching and Referral Hospital in Kenya ^32^. The trial enrolled children aged 6 months to 13 years at admission to hospital if they had complicated SM, defined as: mid-upper arm circumference (MUAC) < 11.5 cm or weight-for-height Z score < −3 if younger than 5 years of age, BMI Z score < −3 if older than 5 years, or oedematous malnutrition at any age. The children were admitted to hospital because of medical complications or failed an appetite test (8/843, 0.9%) as defined by WHO guidelines ^76^. Children were excluded if they had a known allergy to milk products or did not provide consent. The primary outcome of the trial was the time to initial stabilization, defined as having reached the ‘transition’ phase of treatment and switched to a standard higher caloric feed based on WHO guidelines. Biological samples for research including serum and plasma samples were collected upon admission but before randomization and stored at -80°C until analysis. For the trial, biochemical tests were performed to determine serum albumin concentration. Clinical findings were also recorded such as presence of shock, pneumonia, malaria, heart disease, cerebral palsy and diarrhoea, as well as breastfeeding. The trial recruited a total of 843 children of which 8.9% died prior to stabilization while another 6.2% died after the first stabilization ^32^.

#### Validation cohort

The validation cohort was nested within a randomised controlled trial (NCT00934492) that tested the efficacy of daily co-trimoxazole prophylaxis in reducing post-discharge mortality among HIV-uninfected children aged 60 days to 59 months admitted to hospital and diagnosed with SM in four hospitals in Kenya (two rural hospitals in Kilifi and Malindi, and two urban hospitals in Mombasa and Nairobi)) ^33^. Children were eligible for inclusion in the trial on the basis of the mid-upper-arm circumference (MUAC) measurements (<11·5 cm for children aged ≥6 months and <11·0 cm for infants aged 2–5 months) or presence of kwashiorkor; had a negative HIV rapid-antibody test; and had completed the stabilisation phase of treatment. Children were recruited into the trial for a median of 6 days from admission to the hospitals. Children were actively followed up for a total of 1 year, monthly in the first 6 months and every 2 months until month 12 for growth, readmission or death and traced at home if they defaulted. Samples were stored at -80°C until analysis.

### Variables and data source/measurement

The presence of oedema, which was evaluated by trained research clinical staff. Kwashiorkor was diagnosed based on the presence of oedema regardless of concurrent wasting. Children without nutritional oedema and with either mid-upper arm circumference <11.5cm (or <11cm if age <6 months) or weight-for-length/height (WFL/H) < -3 were considered as marasmus.

### Data sources and management

Clinical data was obtained from two independent clinical trials ^32, 33^, as above.

### Study size

Of the 843 study children recruited in the F75 reformulation clinical trial, 46% (385) had grade one (+, n=67), grade two (++, n=113), and grade three (+++, n=29) nutritional oedema at admission. For this study, children with nutritional oedema (n=72; 19%) and with known serum albumin concentrations was selected as the discovery cohort. A total of 181 (21%) had missing albumin concentration at admission. Hence, for the discovery cohort, children with kwashiorkor were matched on serum albumin concentrations to those with marasmus (n=72; 16%). Sample size was limited by finding exact matches of serum albumin concentrations between SM phenotypes and hence all matched case-control pairs were included.

The sample size calculation for the validation cohort was based on the results of the discovery cohort. Based on results for lumican (proteomics) and PC.ae.C34:3 (metabolomics), we calculated that a sample size of 40 per group would be sufficient to achieve >80% power at α = 0.05, accounting for stratification based on oedema severity and multiple testing. Subjects for the validation cohort were selected if they had achieved total nutritional and clinical rehabilitation (defined as having a MUAC >12.5 cm, absence of oedema and/or disease needing hospitalization) at day 60 post-hospital discharge. Selection was also limited to children who had sufficient plasma samples at enrolment and at month 2 of follow-up. Kwashiorkor (n=40) were matched to marasmus (n=40) based on age, sex, and site of recruitment and randomisation arm in the trial.

### Laboratory analyses

#### Untargeted plasma proteomics analysis

Liquid chromatography tandem mass spectrometry plasma proteomics analysis was performed for the discovery cohort using the plasma samples collected at enrolment during admission. Briefly, plasma proteomics was performed using Tandem Mass Tag (TMT; Thermo Scientific) as described in our previous study ^77^. Plasma samples were depleted of abundant proteins using spin columns (Thermo Scientific) then reduced and alkylated respectively with 40mM tris(2-carboxyethyl)phosphine and 80mM iodoacetamide. The proteins were subsequently precipitated using pre-chilled (-20°C) acetone followed by centrifugation at 8,000g for 10 min at 4°C. Precipitated proteins were then subjected to trypsin digestion (1:15 trypsin:sample ratio) and labelled with TMT 10plex kit (Thermo Scientific) according to manufacturer’s instructions. The TMT-tagged peptides generated were then separated on the Dionex Ultimate 3000 nano-flow ultra-high-pressure liquid chromatography system (Thermo Scientific) with a 75 µm × 25 cm C18 reverse-phase analytical column (Thermo Scientific) at 40 °C. Elution was carried out with mobile phase B (80% acetonitrile with 0.1% formic acid) gradient (2 to 35% B) over 310 min at a flow rate of 0.3 μl/min. The chromatographic outflow was connected to a Q Exactive Orbitrap Mass Spectrometer (Thermo Scientific) via a nano-electrospray ion source. Further details of the chromatographic and mass spectrometry parameters were as previously described by Njunge, et al. ^77^.

Mass spectrometer raw files were analysed by MaxQuant software version 1.6.0.1 ^78^ and peptide spectra were searched against the human Uniprot FASTA database using the Andromeda search engine ^79^. Further details on the proteomics dataset extraction and pre-processing were previously in Njunge, et al.^77^. A complete list of all annotated proteins is provided in Supplemental Table S1.

#### Targeted plasma metabolomics analysis

Plasma metabolomics analysis was performed for admission samples only in the discovery cohort using both direct flow injection and reverse-phase liquid chromatography coupled to tandem mass spectrometry (MS/MS) using Prime Platform at The Metabolomics Innovation Centre (University of Alberta, Canada). Mass spectrometric analysis was done using an ABI 4000 Q-Trap mass spectrometer (Applied Biosystems/MDS Sciex, CA, USA), which could provide absolute quantifications of up to 187 endogenous serum metabolites. Moreover, a targeted method for analysis of organic acids from serum was also used to quantify 18 organic acids. A complete list of all targeted metabolites is provided in Supplemental Table S1.

#### Targeted plasma analysis of endothelial glycocalyx components

Plasma levels of Syn1 and HA were performed using quantitative ELISA (Thermo Scientific, MD, USA) following manufacturer’s instructions at admission for the discovery cohort, and at admission and 60 days post-discharge for the validation cohort.

#### Multiplex immunochemical analysis

Magnetic bead-based multiplex assay performed in a Luminex® platform (R&D Systems, MN, USA) were used to quantify plasma concentration of lumican, MMP2, TIMP1, TIMP2. These proteins were assayed at admission for the discovery cohort, and at admission and 60 days post-discharge for the validation cohort. Further, cytokine and chemokines (n = 29) concentration in plasma at admission and 60 days post-discharge for the validation cohort were determined by using a human cytokine magnetic bead assay (EMD Millipore) on the Magpix with Xponent software (version 4.2; Luminex Corp) and acquired Median Fluorescent Intensity data analysed using the Milliplex analyst software (version 3.5.5.0 standard). Levels of CXCL-13, MMP3, 8, 13, and TGF-β were also measured but their concentrations were too low to be detected in most of the samples.

#### Data Preprocessing

For proteomics, columns containing the protein identifiers (IDs), protein names, gene names, and corrected reporter ion intensity in the protein group matrix file from MaxQuant were obtained. Data pre-processing included removing features with more the 20% missing values. The dataset was then log transformed and batch corrected using using *ComBat* function as implemented in *sva* R package ^80^. Missing values were then imputed using an imputation algorithm based on *k*-nearest neighbour.

For metabolomics, metabolites with more than 20% missing values in both SM phenotype groups were removed before analysis. Then, metabolites with concentrations below the limit of detection (LOD) for each metabolite were set as half the LOD. Lastly, only metabolites that have <30% coefficient of variation among the quality control samples were retained for further data analysis. Both proteomics and metabolomics data were log transformed prior to analysis.

#### Data analysis

Data analyses were all performed using R v 3.6 ^81^. Baseline patient characteristics are provided as either mean ± standard deviation (SD), median (25 and 75^th^ percentile) or proportions, as applicable. For the discovery cohort, difference in exposures (untargeted proteome and targeted metabolite levels, Syn1, HA, lumican, MMP2, TIMP1 and TIMP2, and a panel of 29 cytokines and chemokines) between marasmus and kwashiorkor were analysed using conditional logistic regression adjusted for age, sex, HIV status and recruitment site, to account for the sparseness introduced by exactly matching by serum albumin. To assess the association between increasing oedema severity and plasma/serum levels of glycocalyx components, individual protein and metabolite levels, we used an ordinal regression with age, sex, HIV status, recruitment site and serum albumin as additional covariates. High MUAC signifies the absence of wasting but also increases with oedema in kwashiorkor. Hence, no adjustment for MUAC was made in the models as this would obscure the interpretation of the results. Longitudinal analyses were performed using linear mixed models with the individual subjects set as random effect. Correction for multiple testing was performed using Benjamini-Hochberg false-discovery rate method ^82^.

Multi-omics weighted co-expression network analysis (WCNA) was then performed using the WGCNA R package ^83^. WCNA, is a data-driven network approach that is used to find clusters, known as modules, of tightly correlated features that can then be used to assess associations towards a specific outcome^83^. Understanding which modules relate to clinical outcomes and determining pathways involving features that comprise the modules enables us to uncover molecular pathways that could be associated to the outcome of interest. After pre-processing and combining both metabolomics and proteomics datasets, a soft threshold for network construction needed to achieve a scale-free topography typical of biological networks (r² ≳ 0.8)^35^ was determined. Scale-free topography for our data was achieved using β = 18 for an signed network. A Pearson correlation (*s_ij_*) matrix was then generated between each multi-omics pairs (*i* and *j*) which was transformed into an adjacency matrix through the power transformation, *a_ij_* = |*s_ij_*|^β^. This power transformation punishes weak correlations while amplifying strong correlations. Using hierarchical clustering embedded with the WGCNA package, co-expressed metabolites and proteins are clustered into modules. Each member of the module is characterized by an eigenvector (E^(q)^, where (q) denotes the module) through a singular value decomposition. The E^(q)^ represents the collective behaviour of the particular module^35^. To determine which modules are associated with oedematous SAM, a conditional logistic regression was used with E^(q)^ as independent variable adjusted for age, sex, HIV status and site of recruitment stratified by admission serum albumin concentration.

For protein features, the Gene ontology (GO) enriched biological processes (BP) of differentially expressed proteins was determined using The Database for Annotation, Visualization and Integrated Discovery (DAVID) v6.8 Bioinformatics Resource ^84^. Homo sapiens was used as background for enrichment calculation.

#### Data availability and access statement

The processed data and codes (STATA and R) will be deposited in the KEMRI-Wellcome data repository on the Harvard Dataverse under the Biosciences Dataverse subtheme (https://dataverse.harvard.edu/dataverse/kwtrp) and issued with Digital Object Identifiers (DOI) at the time of deposit. The data will be titled “Albumin-dependent and independent mechanisms in the syndrome of kwashiorkor”. The anonymised mass spectrometry raw files generated and analysed in the current study will be deposited to The ProteomeXchange Consortium: http://www.proteomexchange.org/ and assigned a unique identifier.

## Data Availability

The processed data and codes (STATA and R) will be deposited in the KEMRI-Wellcome data repository on the Harvard Dataverse under the Biosciences Dataverse subtheme (https://dataverse.harvard.edu/dataverse/kwtrp) and issued with Digital Object Identifiers (DOI) at the time of deposit. The data will be titled: Albumin-dependent and independent mechanisms in the syndrome of kwashiorkor. The anonymised mass spectrometry raw files generated and analysed in the current study will be deposited to The ProteomeXchange Consortium: http://www.proteomexchange.org/ and assigned a unique identifier.

## Acknowledgement

We thank the parents and guardians of the study participants who patiently participated in in both clinical trials in Malawi and Kenya. We acknowledge the enthusiastic work of our nurses, field workers, clinical and non-clinical staff who tirelessly collected the data and samples and provided administrative support to the project.

GBG was a postdoctoral fellow of the Research Foundation Flanders (FWO) and received financial support from the Thrasher Foundation Early Career Award (15122) and VLIR-UOS-Ghent University Global Minds Fund. JMN, MN, IP, JT, WV, RB, and JAB received support from CHAIN: Bill & Melinda Gates Foundation (OPP1131320). JAB is also supported by MRC/DfID/Wellcome Trust Global Health Trials Scheme (MR/M007367/1). The F75 reformulation trial was funded by Thrasher Research Fund (9403) while the co-trimoxazole trial was funded by the Wellcome Trust (WT083579MA), both awarded to JAB.

## Approvals

The F75 reformulation trial was approved by KEMRI Ethical Review Committee (SCC 2799), College of Medicine Research Ethics Boards of the University of Malawi (P.03/14/1540), Oxford Tropical Research Ethics Committee (OXTREC 58–14) and the Hospital for Sick Children Research Ethics Board, Toronto (1000046559), including the secondary analysis in this manuscript. The co-trimoxazole trials was approved by Kenya National Ethical Review Committee (SSC 1562) and Oxford Tropical Research Ethics Committee (reference number 18-09), including the secondary analysis in this manuscript. This paper is published with the permission of the Director of the Kenya Medical Research Institute.

---

**Supplementary figure S1.**
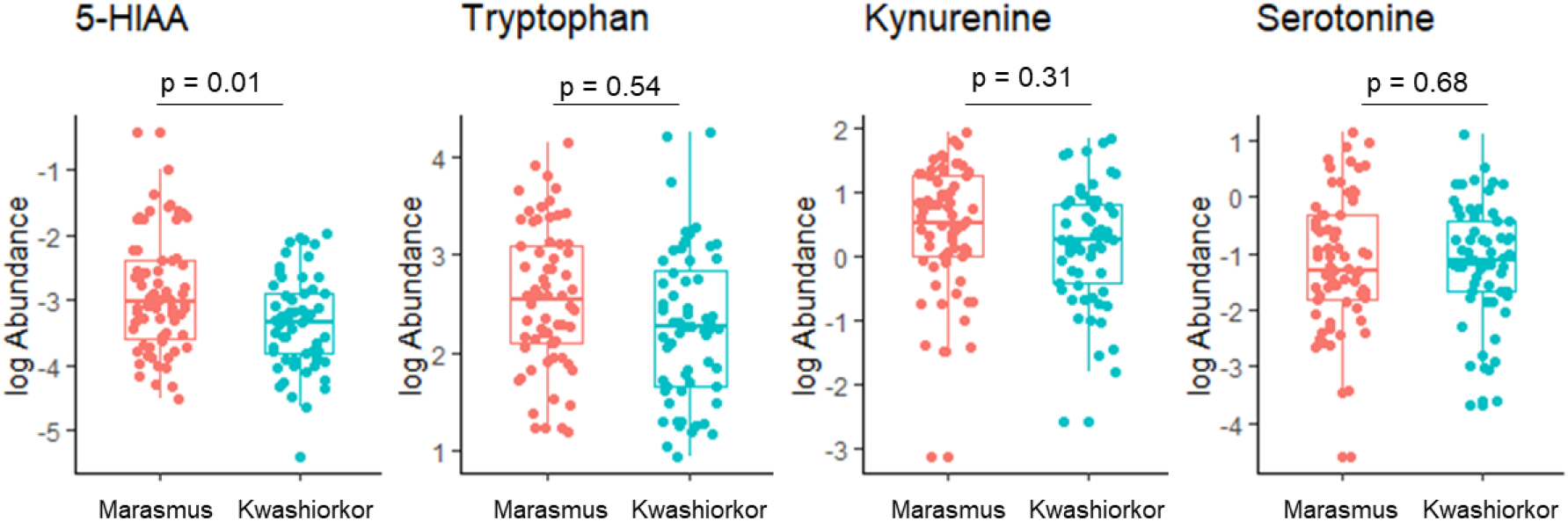
Differences in plasma concentration of tryptophan downstream metabolites between kwashiorkor and marasmus

**Supplementary Table S1.** Individual protein and metabolite associations with kwashiorkor

|  | adjusted odds ratio | std error | pVal | FDR pVal |
| --- | --- | --- | --- | --- |
| *Proteins (Uniprot accession ID)* |  |  |  |  |
| P51884 | 2.72 | 0.25 | 0.00 | 0.01 |
| K7ERI9 | 2.05 | 0.21 | 0.00 | 0.05 |
| P04196 | 2.19 | 0.23 | 0.00 | 0.05 |
| P19823 | 2.11 | 0.23 | 0.00 | 0.05 |
| P19827 | 2.00 | 0.23 | 0.00 | 0.08 |
| Q5VY30 | 2.00 | 0.24 | 0.00 | 0.09 |
| D6RF35 | 1.81 | 0.21 | 0.00 | 0.11 |
| A0A0A0MRJ7 | 1.81 | 0.23 | 0.01 | 0.11 |
| P00738 | 0.58 | 0.21 | 0.01 | 0.11 |
| V9GYM3 | 1.76 | 0.21 | 0.01 | 0.11 |
| P05452 | 1.94 | 0.24 | 0.01 | 0.11 |
| P06396 | 1.86 | 0.23 | 0.01 | 0.11 |
| P29622 | 1.83 | 0.23 | 0.01 | 0.11 |
| P02766 | 1.74 | 0.22 | 0.01 | 0.12 |
| P04004 | 1.72 | 0.22 | 0.01 | 0.12 |
| P04075 | 1.79 | 0.23 | 0.01 | 0.12 |
| P0DJI9 | 0.56 | 0.22 | 0.01 | 0.12 |
| P25311 | 1.78 | 0.22 | 0.01 | 0.12 |
| A0A096LPE2 | 0.58 | 0.22 | 0.01 | 0.12 |
| P01008 | 1.70 | 0.22 | 0.01 | 0.12 |
| P02741 | 0.60 | 0.21 | 0.01 | 0.12 |
| P00915 | 1.75 | 0.23 | 0.02 | 0.13 |
| P02649 | 1.69 | 0.22 | 0.02 | 0.13 |
| P02747 | 1.70 | 0.22 | 0.02 | 0.13 |
| P01042 | 1.66 | 0.22 | 0.02 | 0.14 |
| H0YDW7 | 1.67 | 0.23 | 0.02 | 0.16 |
| A0A087WTK0 | 1.64 | 0.22 | 0.02 | 0.17 |
| A0A087X232 | 1.62 | 0.22 | 0.03 | 0.17 |
| A0A0A0MSV6 | 1.57 | 0.21 | 0.03 | 0.17 |
| I3L145 | 1.60 | 0.21 | 0.03 | 0.17 |
| O75636 | 1.62 | 0.22 | 0.03 | 0.17 |
| P02749 | 1.62 | 0.22 | 0.03 | 0.17 |
| P00747 | 1.57 | 0.21 | 0.03 | 0.18 |
| P02745 | 1.62 | 0.23 | 0.04 | 0.18 |
| P05090 | 1.57 | 0.21 | 0.03 | 0.18 |
| P05546 | 1.65 | 0.24 | 0.03 | 0.18 |
| P69905 | 1.70 | 0.25 | 0.04 | 0.18 |
| P36955 | 1.58 | 0.22 | 0.04 | 0.19 |
| P43251 | 1.57 | 0.22 | 0.04 | 0.19 |
| P68871 | 1.67 | 0.25 | 0.04 | 0.19 |
| Q16610 | 1.55 | 0.22 | 0.04 | 0.19 |
| B4DPQ0 | 1.51 | 0.21 | 0.05 | 0.20 |
| I3LOA1 | 1.53 | 0.22 | 0.05 | 0.20 |
| P00918 | 1.55 | 0.23 | 0.05 | 0.20 |
| P01023 | 1.56 | 0.23 | 0.05 | 0.20 |
| P02751 | 1.52 | 0.21 | 0.05 | 0.20 |
| P02765 | 1.53 | 0.22 | 0.05 | 0.20 |
| P06727 | 1.55 | 0.23 | 0.05 | 0.20 |
| Q08380 | 1.54 | 0.22 | 0.05 | 0.20 |
| P05155 | 1.50 | 0.21 | 0.06 | 0.21 |
| P22792 | 1.51 | 0.22 | 0.06 | 0.21 |
| P01024 | 1.48 | 0.21 | 0.06 | 0.21 |
| P07225 | 1.53 | 0.23 | 0.06 | 0.21 |
| P41222 | 1.48 | 0.21 | 0.06 | 0.21 |
| P02042 | 1.56 | 0.24 | 0.07 | 0.22 |
| P05109 | 0.67 | 0.22 | 0.07 | 0.22 |
| P48740 | 1.49 | 0.22 | 0.07 | 0.22 |
| P02743 | 1.46 | 0.21 | 0.07 | 0.22 |
| P27918 | 1.48 | 0.22 | 0.07 | 0.22 |
| P43652 | 1.47 | 0.22 | 0.07 | 0.22 |
| P33151 | 1.46 | 0.21 | 0.08 | 0.23 |
| A0A1W2PQB1 | 1.39 | 0.19 | 0.08 | 0.23 |
| P02760 | 1.47 | 0.22 | 0.08 | 0.23 |
| P08697 | 1.47 | 0.22 | 0.08 | 0.23 |
| P0C0L4 | 1.47 | 0.22 | 0.08 | 0.23 |
| Q9NZP8 | 1.47 | 0.22 | 0.08 | 0.23 |
| P55056 | 1.42 | 0.20 | 0.08 | 0.23 |
| P04180 | 1.41 | 0.20 | 0.09 | 0.23 |
| P08603 | 1.43 | 0.21 | 0.09 | 0.23 |
| Q96PD5 | 1.43 | 0.21 | 0.09 | 0.23 |
| P02750 | 1.43 | 0.21 | 0.09 | 0.24 |
| P08571 | 1.42 | 0.21 | 0.10 | 0.24 |
| P0DJI8 | 0.71 | 0.21 | 0.09 | 0.24 |
| P32119 | 1.46 | 0.23 | 0.09 | 0.24 |
| Q14520 | 1.42 | 0.21 | 0.10 | 0.24 |
| P04217 | 1.44 | 0.22 | 0.10 | 0.24 |
| P00742 | 1.44 | 0.22 | 0.10 | 0.24 |
| P08185 | 1.42 | 0.22 | 0.10 | 0.24 |
| J3QQX6 | 1.41 | 0.21 | 0.10 | 0.24 |
| P02774 | 1.41 | 0.21 | 0.10 | 0.24 |
| H0Y2X5 | 1.39 | 0.21 | 0.11 | 0.24 |
| P07360 | 1.41 | 0.22 | 0.11 | 0.24 |
| P21333 | 1.39 | 0.20 | 0.11 | 0.24 |
| G3XAK1 | 1.39 | 0.21 | 0.11 | 0.25 |
| P05543 | 1.40 | 0.21 | 0.11 | 0.25 |
| Q6UXB8 | 1.40 | 0.21 | 0.12 | 0.25 |
| P06702 | 0.70 | 0.22 | 0.12 | 0.25 |
| G3XAM2 | 1.40 | 0.22 | 0.12 | 0.26 |
| P10909 | 1.43 | 0.23 | 0.12 | 0.26 |
| P27169 | 1.38 | 0.21 | 0.12 | 0.26 |
| P04264 | 1.38 | 0.21 | 0.13 | 0.26 |
| O95445 | 1.36 | 0.20 | 0.13 | 0.27 |
| CON__ENSEMBL_ENSBTAP00000024466 | 1.37 | 0.21 | 0.14 | 0.28 |
| E7END6 | 1.35 | 0.20 | 0.14 | 0.28 |
| P00734 | 1.36 | 0.22 | 0.15 | 0.29 |
| F5GZZ9 | 1.35 | 0.21 | 0.15 | 0.29 |
| P04114 | 1.34 | 0.21 | 0.16 | 0.31 |
| H0YAC1 | 1.35 | 0.22 | 0.17 | 0.32 |
| P0C0L5 | 1.35 | 0.22 | 0.17 | 0.32 |
| A0A0A0MS08 | 1.32 | 0.21 | 0.18 | 0.33 |
| P00748 | 1.29 | 0.20 | 0.20 | 0.36 |
| P14151 | 1.30 | 0.21 | 0.20 | 0.36 |
| V9GYE7 | 1.29 | 0.20 | 0.20 | 0.36 |
| Q13201 | 1.30 | 0.21 | 0.20 | 0.36 |
| C9JXF9 | 1.28 | 0.20 | 0.21 | 0.37 |
| P00450 | 1.31 | 0.22 | 0.21 | 0.37 |
| P10643 | 1.29 | 0.21 | 0.22 | 0.37 |
| Q03591 | 1.28 | 0.20 | 0.21 | 0.37 |
| P22692 | 1.27 | 0.20 | 0.23 | 0.39 |
| Q04756 | 1.31 | 0.22 | 0.23 | 0.39 |
| B4E1Z4 | 1.27 | 0.20 | 0.23 | 0.39 |
| K7ERG9 | 1.26 | 0.20 | 0.25 | 0.41 |
| B0YIW2 | 1.26 | 0.21 | 0.26 | 0.43 |
| P02775 | 1.27 | 0.22 | 0.27 | 0.43 |
| H7BYH4 | 1.26 | 0.21 | 0.27 | 0.43 |
| P02671 | 0.79 | 0.21 | 0.27 | 0.43 |
| P15169 | 1.25 | 0.21 | 0.27 | 0.43 |
| A0A286YFY4 | 1.23 | 0.19 | 0.29 | 0.45 |
| P05362 | 1.25 | 0.21 | 0.29 | 0.45 |
| CON__P02768_1 | 1.25 | 0.21 | 0.30 | 0.46 |
| A0A024ROT9 | 1.23 | 0.20 | 0.30 | 0.46 |
| P13671 | 1.24 | 0.21 | 0.31 | 0.47 |
| P10451 | 1.23 | 0.21 | 0.32 | 0.48 |
| P07996 | 1.24 | 0.22 | 0.32 | 0.48 |
| P02776 | 1.25 | 0.23 | 0.33 | 0.49 |
| P01706 | 1.22 | 0.21 | 0.34 | 0.49 |
| P62937 | 1.24 | 0.22 | 0.33 | 0.49 |
| Q12805 | 1.22 | 0.21 | 0.34 | 0.49 |
| P02675 | 0.82 | 0.21 | 0.35 | 0.51 |
| P02763 | 1.22 | 0.22 | 0.36 | 0.51 |
| CON__Q1RMN8 | 1.20 | 0.20 | 0.36 | 0.51 |
| P02790 | 1.19 | 0.20 | 0.37 | 0.51 |
| P80748 | 0.83 | 0.22 | 0.38 | 0.53 |
| P13796 | 1.18 | 0.20 | 0.39 | 0.54 |
| P02787 | 1.19 | 0.21 | 0.43 | 0.58 |
| P06681 | 1.16 | 0.19 | 0.43 | 0.58 |
| Q06033 | 1.18 | 0.21 | 0.43 | 0.58 |
| P00740 | 1.17 | 0.20 | 0.44 | 0.59 |
| P04275 | 0.85 | 0.21 | 0.44 | 0.59 |
| O75882 | 1.17 | 0.21 | 0.45 | 0.59 |
| P04003 | 1.17 | 0.21 | 0.45 | 0.59 |
| A0A286YFY1 | 0.85 | 0.23 | 0.48 | 0.61 |
| CON__P00761 | 1.16 | 0.21 | 0.47 | 0.61 |
| P02679 | 0.86 | 0.21 | 0.47 | 0.61 |
| P23142 | 1.15 | 0.20 | 0.48 | 0.62 |
| P60709 | 1.16 | 0.21 | 0.49 | 0.62 |
| O00391 | 0.87 | 0.20 | 0.49 | 0.62 |
| Q15582 | 1.15 | 0.21 | 0.49 | 0.62 |
| A0A2Q2TTZ9 | 1.17 | 0.23 | 0.50 | 0.62 |
| P62328 | 0.87 | 0.21 | 0.51 | 0.63 |
| P00739 | 0.88 | 0.20 | 0.52 | 0.64 |
| P01780 | 1.14 | 0.21 | 0.54 | 0.66 |
| P01834 | 1.13 | 0.21 | 0.57 | 0.69 |
| P01871 | 0.90 | 0.20 | 0.58 | 0.70 |
| G3XAP6 | 1.11 | 0.20 | 0.59 | 0.71 |
| P01031 | 1.12 | 0.21 | 0.61 | 0.72 |
| P01011 | 0.91 | 0.21 | 0.64 | 0.75 |
| P01619 | 1.10 | 0.21 | 0.64 | 0.75 |
| P20851 | 1.10 | 0.21 | 0.64 | 0.75 |
| P07357 | 1.09 | 0.20 | 0.66 | 0.76 |
| P18065 | 0.92 | 0.20 | 0.66 | 0.76 |
| P0DOY3 | 1.09 | 0.22 | 0.71 | 0.81 |
| A0A087X0M8 | 1.08 | 0.20 | 0.71 | 0.81 |
| A0A2R8Y7X9 | 0.92 | 0.22 | 0.71 | 0.81 |
| P19320 | 1.08 | 0.21 | 0.72 | 0.81 |
| A0A0A0MRZ8 | 1.07 | 0.20 | 0.76 | 0.85 |
| P60174 | 1.06 | 0.21 | 0.77 | 0.85 |
| D6RGE2 | 1.06 | 0.22 | 0.78 | 0.86 |
| P07358 | 1.06 | 0.20 | 0.79 | 0.86 |
| S4R460 | 0.95 | 0.20 | 0.79 | 0.87 |
| O43866 | 0.95 | 0.20 | 0.80 | 0.87 |
| P61769 | 1.05 | 0.21 | 0.81 | 0.87 |
| A0A0A0MSA9 | 0.96 | 0.22 | 0.84 | 0.90 |
| P04040 | 1.05 | 0.23 | 0.85 | 0.90 |
| A0A0U1RR20 | 1.03 | 0.20 | 0.88 | 0.94 |
| A0A024R6I7 | 1.02 | 0.19 | 0.92 | 0.96 |
| E9PFZ2 | 1.02 | 0.20 | 0.93 | 0.96 |
| P01034 | 1.02 | 0.20 | 0.93 | 0.96 |
| P01591 | 0.98 | 0.21 | 0.93 | 0.96 |
| A0A286YES1 | 0.98 | 0.21 | 0.94 | 0.97 |
| O14791 | 0.98 | 0.21 | 0.94 | 0.97 |
| P01019 | 1.01 | 0.21 | 0.96 | 0.98 |
| P02748 | 1.01 | 0.20 | 0.97 | 0.99 |
| P19652 | 1.01 | 0.21 | 0.98 | 0.99 |
| Q14624 | 1.00 | 0.20 | 0.99 | 0.99 |
| A0A0B4J231 | 1.00 | 0.21 | 1.00 | 1.00 |
| *Metabolites* |  |  |  |  |
| AA_Ala | 1.25 | 0.22 | 0.32 | 0.64 |
| AA_Asn | 0.80 | 0.20 | 0.27 | 0.61 |
| AA_Asp | 0.82 | 0.24 | 0.42 | 0.68 |
| AA_Cit | 1.11 | 0.20 | 0.60 | 0.77 |
| AA_Gln | 1.36 | 0.22 | 0.17 | 0.46 |
| AA_Gly | 1.47 | 0.24 | 0.10 | 0.34 |
| AA_His | 1.01 | 0.21 | 0.96 | 0.97 |
| AA_Ile | 0.83 | 0.22 | 0.40 | 0.68 |
| AA_Leu | 0.67 | 0.23 | 0.08 | 0.31 |
| AA_Lys | 0.65 | 0.26 | 0.11 | 0.34 |
| AA_Met | 0.67 | 0.24 | 0.10 | 0.34 |
| AA_Orn | 0.78 | 0.23 | 0.29 | 0.62 |
| AA_Phe | 0.47 | 0.25 | 0.00 | 0.04 |
| AA_Pro | 1.38 | 0.24 | 0.18 | 0.48 |
| AA_Thr | 0.80 | 0.21 | 0.28 | 0.62 |
| AA_Trp | 0.72 | 0.26 | 0.21 | 0.54 |
| AA_Tyr | 0.62 | 0.25 | 0.05 | 0.28 |
| AA_Val | 0.67 | 0.25 | 0.10 | 0.34 |
| AC_C0 | 0.79 | 0.20 | 0.24 | 0.59 |
| AC_C10 | 0.65 | 0.22 | 0.06 | 0.28 |
| AC_C12 | 0.94 | 0.22 | 0.79 | 0.88 |
| AC_C14.1 | 1.22 | 0.21 | 0.35 | 0.68 |
| AC_C16.1 | 1.55 | 0.23 | 0.06 | 0.28 |
| AC_C16.2 | 1.95 | 0.30 | 0.02 | 0.19 |
| AC_C18 | 1.57 | 0.24 | 0.06 | 0.28 |
| AC_C18.1 | 1.51 | 0.24 | 0.08 | 0.31 |
| AC_C18.2 | 1.73 | 0.30 | 0.06 | 0.28 |
| AC_C2 | 0.61 | 0.24 | 0.04 | 0.23 |
| AC_C3 | 0.80 | 0.22 | 0.32 | 0.64 |
| AC_C3.DC..C4.OH. | 1.17 | 0.21 | 0.47 | 0.68 |
| AC_C4 | 0.66 | 0.23 | 0.08 | 0.31 |
| AC_C5 | 0.35 | 0.50 | 0.04 | 0.22 |
| AC_C5.DC..C6.OH. | 0.75 | 0.25 | 0.25 | 0.60 |
| AC_C9 | 1.22 | 0.25 | 0.41 | 0.68 |
| BA_Ac.Orn | 0.88 | 0.23 | 0.58 | 0.76 |
| BA_alpha.AAA | 0.69 | 0.24 | 0.13 | 0.38 |
| BA_Creatinine | 0.57 | 0.26 | 0.03 | 0.22 |
| BA_Kynurenine | 0.66 | 0.24 | 0.08 | 0.31 |
| BA_Met.SO | 0.71 | 0.31 | 0.27 | 0.61 |
| BA_Putrescine | 1.16 | 0.21 | 0.48 | 0.68 |
| BA_Serotonin | 0.82 | 0.23 | 0.39 | 0.68 |
| BA_Spermidine | 1.45 | 0.23 | 0.11 | 0.35 |
| BA_Spermine | 1.04 | 0.22 | 0.85 | 0.91 |
| BA_t4.OH.Pro | 0.68 | 0.27 | 0.16 | 0.45 |
| BA_Taurine | 1.03 | 0.22 | 0.89 | 0.94 |
| GP_lysoPC.a.C16.0 | 2.06 | 0.29 | 0.01 | 0.12 |
| GP_lysoPC.a.C16.1 | 3.51 | 0.33 | 0.00 | 0.01 |
| GP_lysoPC.a.C17.0 | 1.70 | 0.28 | 0.06 | 0.28 |
| GP_lysoPC.a.C18.0 | 3.11 | 0.35 | 0.00 | 0.02 |
| GP_lysoPC.a.C18.1 | 3.43 | 0.33 | 0.00 | 0.01 |
| GP_lysoPC.a.C18.2 | 5.53 | 0.43 | 0.00 | 0.01 |
| GP_lysoPC.a.C20.3 | 3.81 | 0.39 | 0.00 | 0.02 |
| GP_lysoPC.a.C20.4 | 3.87 | 0.36 | 0.00 | 0.01 |
| GP_lysoPC.a.C28.0 | 0.85 | 0.21 | 0.43 | 0.68 |
| GP_PC.aa.C28.1 | 1.05 | 0.22 | 0.83 | 0.90 |
| GP_PC.aa.C30.0 | 0.81 | 0.24 | 0.39 | 0.68 |
| GP_PC.aa.C32.0 | 0.80 | 0.22 | 0.32 | 0.64 |
| GP_PC.aa.C32.1 | 0.94 | 0.24 | 0.80 | 0.88 |
| GP_PC.aa.C32.2 | 1.22 | 0.21 | 0.36 | 0.68 |
| GP_PC.aa.C32.3 | 1.89 | 0.25 | 0.01 | 0.11 |
| GP_PC.aa.C34.1 | 0.76 | 0.26 | 0.28 | 0.62 |
| GP_PC.aa.C34.2 | 1.17 | 0.20 | 0.43 | 0.68 |
| GP_PC.aa.C34.3 | 1.71 | 0.23 | 0.02 | 0.18 |
| GP_PC.aa.C34.4 | 1.49 | 0.24 | 0.09 | 0.32 |
| GP_PC.aa.C36.0 | 0.84 | 0.24 | 0.46 | 0.68 |
| GP_PC.aa.C36.1 | 1.20 | 0.21 | 0.40 | 0.68 |
| GP_PC.aa.C36.2 | 1.68 | 0.23 | 0.02 | 0.18 |
| GP_PC.aa.C36.3 | 1.56 | 0.23 | 0.06 | 0.28 |
| GP_PC.aa.C36.4 | 1.11 | 0.20 | 0.60 | 0.77 |
| GP_PC.aa.C36.5 | 1.04 | 0.22 | 0.85 | 0.91 |
| GP_PC.aa.C36.6 | 0.95 | 0.24 | 0.82 | 0.90 |
| GP_PC.aa.C38.0 | 0.88 | 0.20 | 0.55 | 0.74 |
| GP_PC.aa.C38.3 | 1.54 | 0.26 | 0.10 | 0.34 |
| GP_PC.aa.C38.4 | 1.25 | 0.22 | 0.33 | 0.64 |
| GP_PC.aa.C38.5 | 1.20 | 0.22 | 0.42 | 0.68 |
| GP_PC.aa.C38.6 | 0.80 | 0.23 | 0.33 | 0.64 |
| GP_PC.aa.C40.1 | 0.86 | 0.21 | 0.48 | 0.68 |
| GP_PC.aa.C40.2 | 0.86 | 0.22 | 0.47 | 0.68 |
| GP_PC.aa.C40.3 | 1.06 | 0.21 | 0.77 | 0.87 |
| GP_PC.aa.C40.4 | 1.23 | 0.23 | 0.37 | 0.68 |
| GP_PC.aa.C40.5 | 1.18 | 0.22 | 0.46 | 0.68 |
| GP_PC.aa.C40.6 | 0.94 | 0.22 | 0.79 | 0.88 |
| GP_PC.aa.C42.0 | 0.86 | 0.22 | 0.51 | 0.71 |
| GP_PC.aa.C42.1 | 0.89 | 0.21 | 0.56 | 0.75 |
| GP_PC.aa.C42.2 | 0.94 | 0.19 | 0.75 | 0.87 |
| GP_PC.aa.C42.4 | 1.26 | 0.23 | 0.31 | 0.64 |
| GP_PC.aa.C42.5 | 1.63 | 0.24 | 0.05 | 0.26 |
| GP_PC.aa.C42.6 | 1.23 | 0.21 | 0.33 | 0.64 |
| GP_PC.ae.C30.0 | 0.83 | 0.23 | 0.42 | 0.68 |
| GP_PC.ae.C32.1 | 1.13 | 0.21 | 0.56 | 0.75 |
| GP_PC.ae.C32.2 | 1.67 | 0.24 | 0.03 | 0.21 |
| GP_PC.ae.C34.0 | 0.93 | 0.21 | 0.71 | 0.86 |
| GP_PC.ae.C34.1 | 1.13 | 0.21 | 0.56 | 0.75 |
| GP_PC.ae.C34.2 | 1.83 | 0.23 | 0.01 | 0.10 |
| GP_PC.ae.C34.3 | 4.55 | 0.36 | 0.00 | 0.00 |
| GP_PC.ae.C36.0 | 0.59 | 0.30 | 0.08 | 0.31 |
| GP_PC.ae.C36.1 | 0.90 | 0.20 | 0.59 | 0.76 |
| GP_PC.ae.C36.2 | 1.27 | 0.21 | 0.25 | 0.61 |
| GP_PC.ae.C36.3 | 1.66 | 0.23 | 0.03 | 0.21 |
| GP_PC.ae.C36.4 | 1.22 | 0.22 | 0.38 | 0.68 |
| GP_PC.ae.C36.5 | 1.82 | 0.23 | 0.01 | 0.11 |
| GP_PC.ae.C38.0 | 1.06 | 0.20 | 0.76 | 0.87 |
| GP_PC.ae.C38.2 | 0.99 | 0.18 | 0.98 | 0.98 |
| GP_PC.ae.C38.3 | 1.08 | 0.21 | 0.72 | 0.86 |
| GP_PC.ae.C38.4 | 1.26 | 0.22 | 0.28 | 0.62 |
| GP_PC.ae.C38.5 | 1.43 | 0.23 | 0.13 | 0.38 |
| GP_PC.ae.C38.6 | 1.21 | 0.21 | 0.36 | 0.68 |
| GP_PC.ae.C40.1 | 1.16 | 0.24 | 0.53 | 0.73 |
| GP_PC.ae.C40.2 | 0.98 | 0.20 | 0.91 | 0.94 |
| GP_PC.ae.C40.3 | 1.11 | 0.21 | 0.62 | 0.77 |
| GP_PC.ae.C40.4 | 1.18 | 0.22 | 0.45 | 0.68 |
| GP_PC.ae.C40.5 | 1.33 | 0.22 | 0.19 | 0.50 |
| GP_PC.ae.C40.6 | 1.11 | 0.21 | 0.61 | 0.77 |
| GP_PC.ae.C42.1 | 0.96 | 0.19 | 0.85 | 0.91 |
| GP_PC.ae.C42.2 | 0.91 | 0.21 | 0.63 | 0.77 |
| GP_PC.ae.C42.3 | 0.94 | 0.21 | 0.77 | 0.87 |
| GP_PC.ae.C42.4 | 0.91 | 0.23 | 0.70 | 0.84 |
| GP_PC.ae.C42.5 | 1.19 | 0.22 | 0.44 | 0.68 |
| GP_PC.ae.C44.3 | 0.82 | 0.21 | 0.33 | 0.64 |
| GP_PC.ae.C44.4 | 0.92 | 0.27 | 0.75 | 0.87 |
| GP_PC.ae.C44.5 | 0.81 | 0.26 | 0.41 | 0.68 |
| GP_PC.ae.C44.6 | 0.92 | 0.22 | 0.69 | 0.84 |
| Hexose | 0.68 | 0.25 | 0.11 | 0.35 |
| OA_5.Hydroxyindole.acetic.acid | 0.19 | 0.47 | 0.00 | 0.01 |
| OA_alpha.Ketoglutaric.acid | 1.03 | 0.22 | 0.91 | 0.94 |
| OA_beta.Hydroxybutyric.acid | 0.79 | 0.21 | 0.27 | 0.61 |
| OA_Butyric.acid | 0.98 | 0.22 | 0.94 | 0.96 |
| OA_Citric.acid | 0.90 | 0.21 | 0.61 | 0.77 |
| OA_Fumaric.acid | 1.00 | 0.21 | 1.00 | 1.00 |
| OA_Hippuric.acid | 0.66 | 0.24 | 0.08 | 0.31 |
| OA_Homovanillic.acid | 0.44 | 0.56 | 0.14 | 0.41 |
| OA_HPHPA | 3.65 | 0.66 | 0.05 | 0.28 |
| OA_Indole.acetic.acid | 1.14 | 0.19 | 0.48 | 0.68 |
| OA_Isobutyric.acid | 0.37 | 0.40 | 0.01 | 0.12 |
| OA_Lactic.acid | 1.03 | 0.23 | 0.91 | 0.94 |
| OA_Methylmalonic.acid | 0.74 | 0.21 | 0.16 | 0.45 |
| OA_p.Hydroxyphenylacetic.acid | 0.86 | 0.20 | 0.46 | 0.68 |
| OA_Propionic.acid | 1.01 | 0.19 | 0.94 | 0.96 |
| OA_Pyruvic.acid | 1.07 | 0.20 | 0.74 | 0.87 |
| OA_Succinic.acid | 0.85 | 0.21 | 0.42 | 0.68 |
| OA_Uric.acid | 0.38 | 0.39 | 0.01 | 0.12 |
| SL_SM.OH..C14.1 | 1.15 | 0.22 | 0.52 | 0.73 |
| SL_SM.OH..C16.1 | 1.28 | 0.22 | 0.26 | 0.61 |
| SL_SM.OH..C22.1 | 1.08 | 0.21 | 0.72 | 0.86 |
| SL_SM.OH..C22.2 | 1.39 | 0.22 | 0.14 | 0.40 |
| SL_SM.OH..C24.1 | 1.17 | 0.22 | 0.47 | 0.68 |
| SL_SM.C16.0 | 1.14 | 0.22 | 0.54 | 0.74 |
| SL_SM.C16.1 | 1.70 | 0.25 | 0.03 | 0.21 |
| SL_SM.C18.0 | 1.49 | 0.22 | 0.07 | 0.28 |
| SL_SM.C18.1 | 2.22 | 0.25 | 0.00 | 0.02 |
| SL_SM.C20.2 | 2.26 | 0.29 | 0.00 | 0.07 |
| SL_SM.C24.0 | 1.32 | 0.23 | 0.23 | 0.58 |
| SL_SM.C24.1 | 1.20 | 0.23 | 0.42 | 0.68 |
| SL_SM.C26.0 | 1.02 | 0.21 | 0.90 | 0.94 |
| SL_SM.C26.1 | 1.11 | 0.21 | 0.62 | 0.77 |

**Supplementary Table S2.**
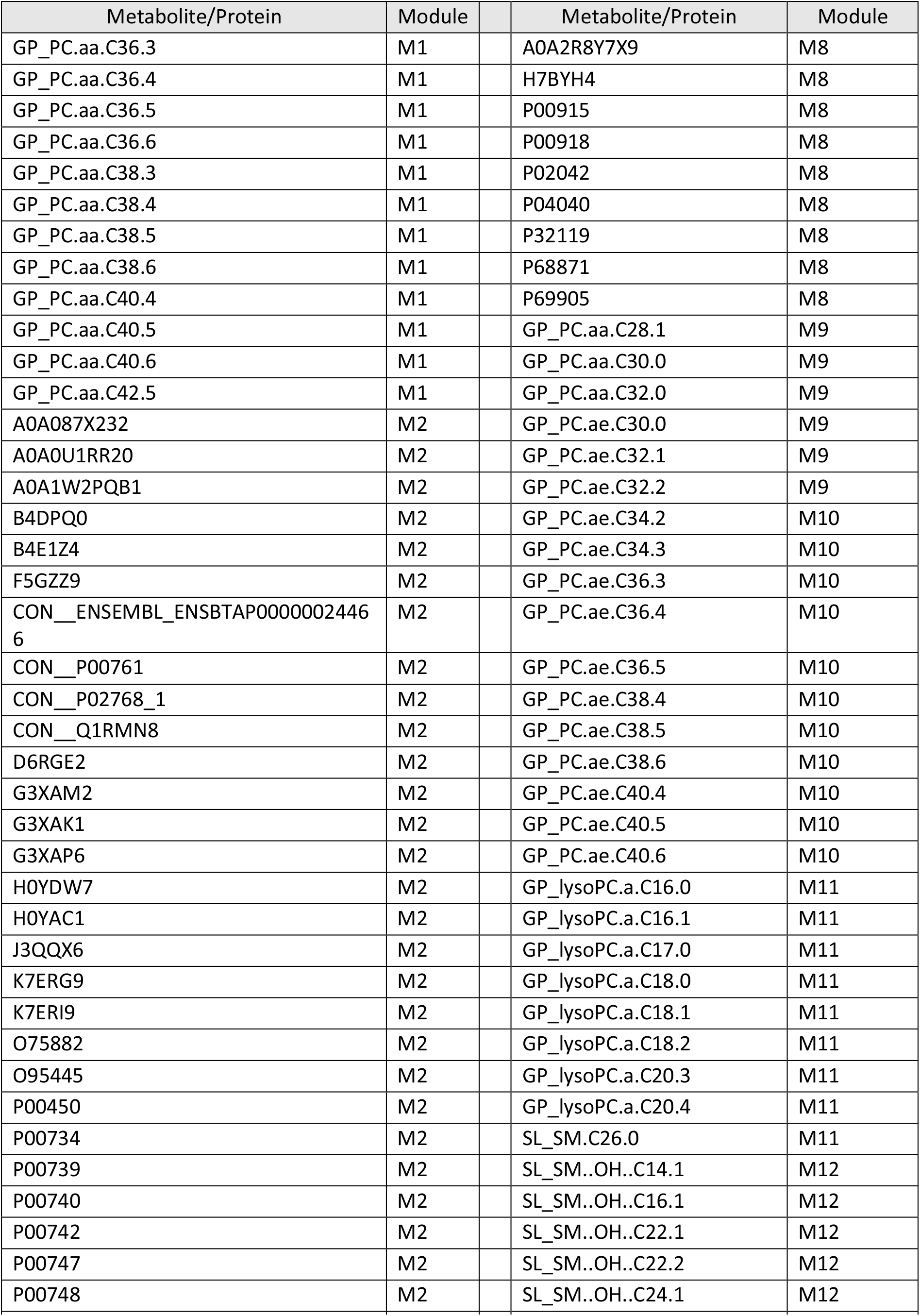
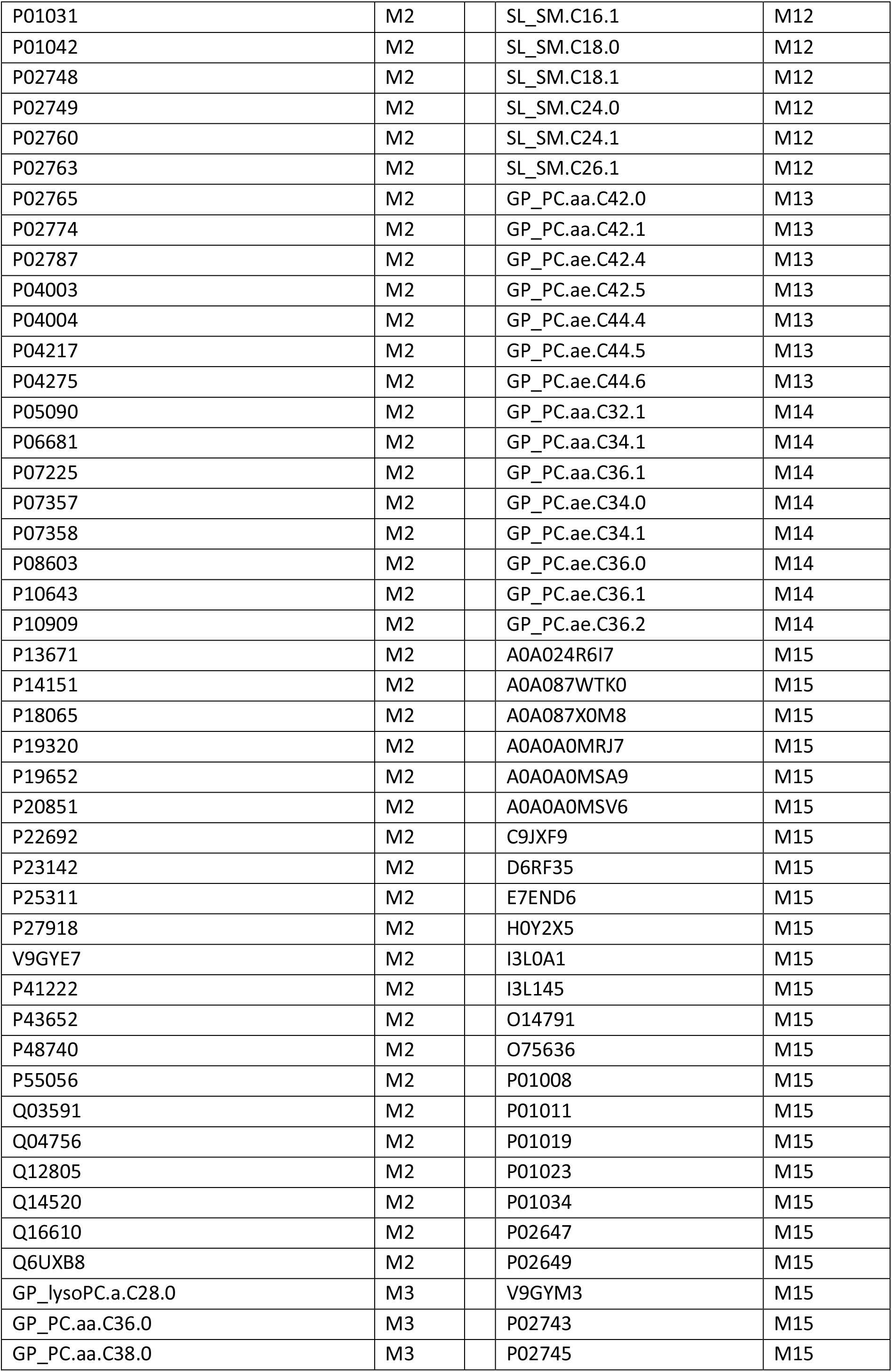
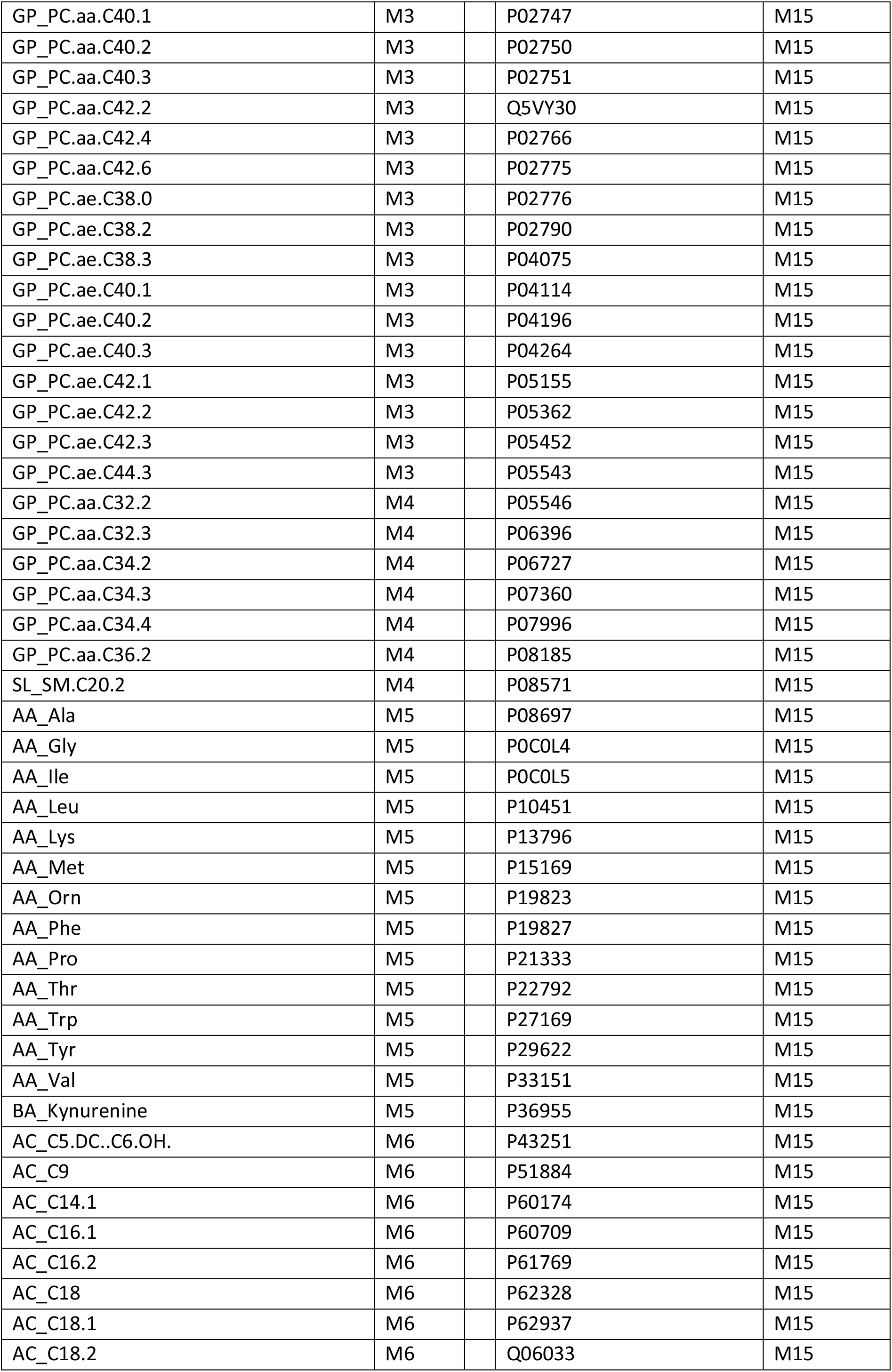
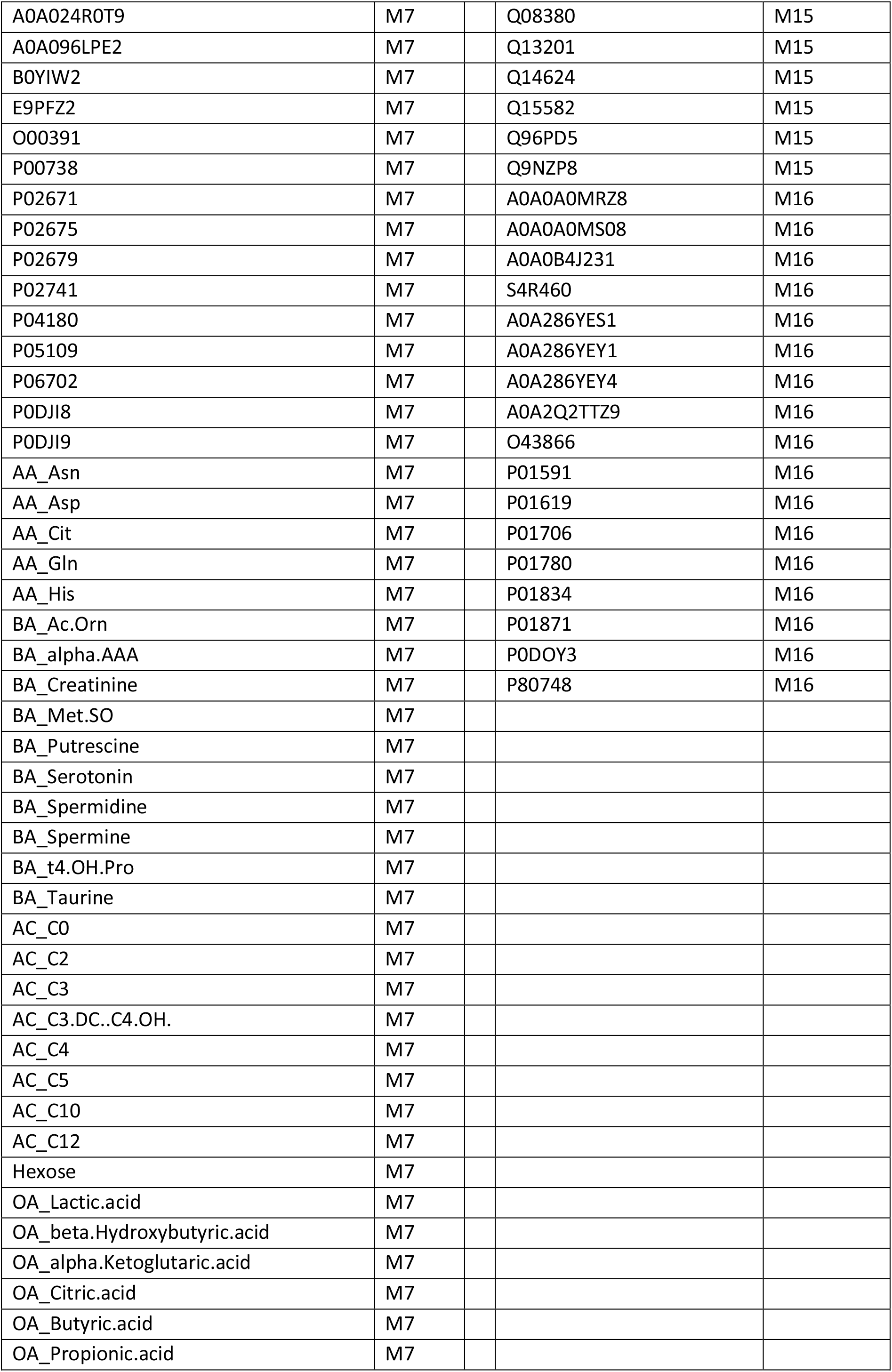
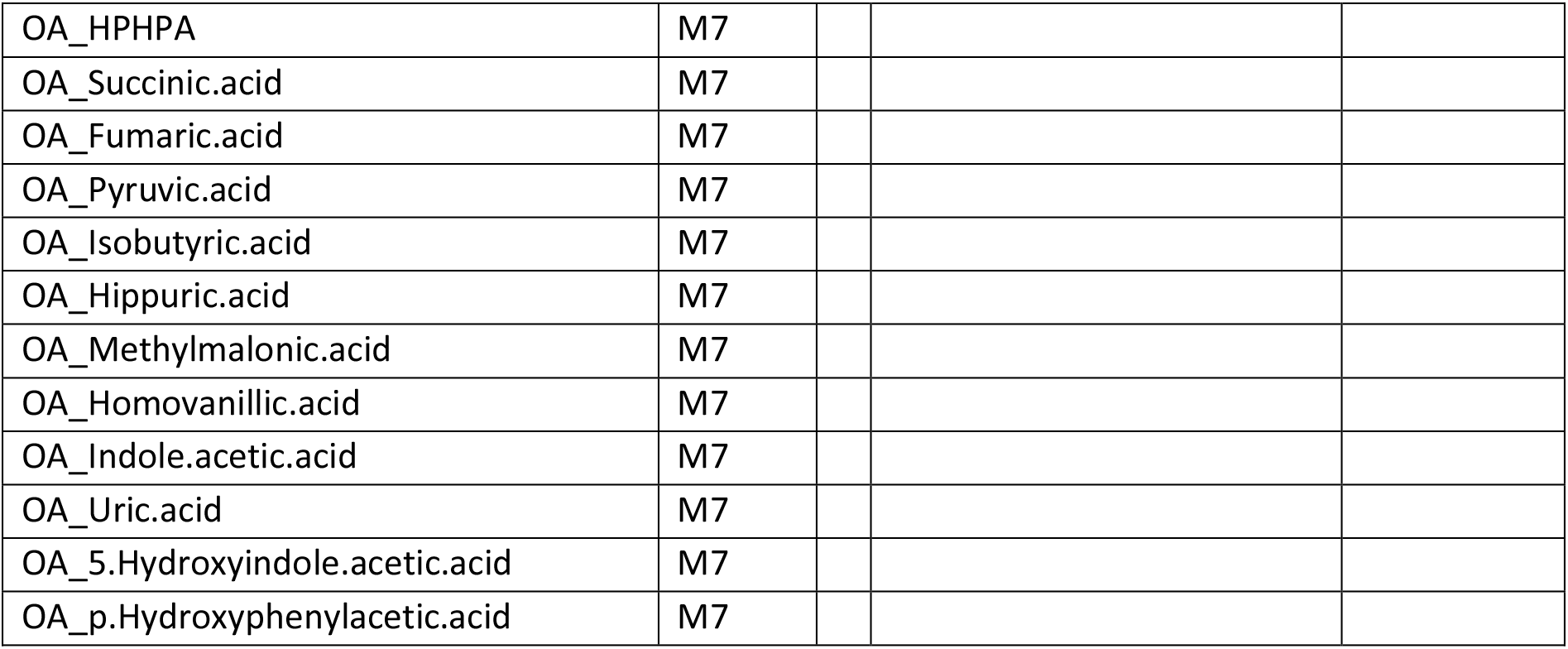
Membership of proteins and metabolites to modules in the weighted correlation network analysis

| Metabolite/Protein | Module | Metabolite/Protein | Module |
| --- | --- | --- | --- |
| GP_PC.aa.C36.3 | M1 | A0A2R8Y7X9 | M8 |
| GP_PC.aa.C36.4 | M1 | H7BYH4 | M8 |
| GP_PC.aa.C36.5 | M1 | P00915 | M8 |
| GP_PC.aa.C36.6 | M1 | P00918 | M8 |
| GP_PC.aa.C38.3 | M1 | P02042 | M8 |
| GP_PC.aa.C38.4 | M1 | P04040 | M8 |
| GP_PC.aa.C38.5 | M1 | P32119 | M8 |
| GP_PC.aa.C38.6 | M1 | P68871 | M8 |
| GP_PC.aa.C40.4 | M1 | P69905 | M8 |
| GP_PC.aa.C40.5 | M1 | GP_PC.aa.C28.1 | M9 |
| GP_PC.aa.C40.6 | M1 | GP_PC.aa.C30.0 | M9 |
| GP_PC.aa.C42.5 | M1 | GP_PC.aa.C32.0 | M9 |
| A0A087X232 | M2 | GP_PC.ae.C30.0 | M9 |
| A0A0U1RR20 | M2 | GP_PC.ae.C32.1 | M9 |
| A0A1W2PQB1 | M2 | GP_PC.ae.C32.2 | M9 |
| B4DPQ0 | M2 | GP_PC.ae.C34.2 | M10 |
| B4E1Z4 | M2 | GP_PC.ae.C34.3 | M10 |
| F5GZZ9 | M2 | GP_PC.ae.C36.3 | M10 |
| CON__ENSEMBL_ENSBTAP00000024466 | M2 | GP_PC.ae.C36.4 | M10 |
| CON__P00761 | M2 | GP_PC.ae.C36.5 | M10 |
| CON__P02768_1 | M2 | GP_PC.ae.C38.4 | M10 |
| CON__Q1RMN8 | M2 | GP_PC.ae.C38.5 | M10 |
| D6RGE2 | M2 | GP_PC.ae.C38.6 | M10 |
| G3XAM2 | M2 | GP_PC.ae.C40.4 | M10 |
| G3XAK1 | M2 | GP_PC.ae.C40.5 | M10 |
| G3XAP6 | M2 | GP_PC.ae.C40.6 | M10 |
| H0YDW7 | M2 | GP_lysoPC.a.C16.0 | M11 |
| H0YAC1 | M2 | GP_lysoPC.a.C16.1 | M11 |
| J3QQX6 | M2 | GP_lysoPC.a.C17.0 | M11 |
| K7ERG9 | M2 | GP_lysoPC.a.C18.0 | M11 |
| K7ERI9 | M2 | GP_lysoPC.a.C18.1 | M11 |
| O75882 | M2 | GP_lysoPC.a.C18.2 | M11 |
| O95445 | M2 | GP_lysoPC.a.C20.3 | M11 |
| P00450 | M2 | GP_lysoPC.a.C20.4 | M11 |
| P00734 | M2 | SL_SM.C26.0 | M11 |
| P00739 | M2 | SL_SM.OH.C14.1 | M12 |
| P00740 | M2 | SL_SM.OH.C16.1 | M12 |
| P00742 | M2 | SL_SM.OH.C22.1 | M12 |
| P00747 | M2 | SL_SM.OH.C22.2 | M12 |
| P00748 | M2 | SL_SM.OH.C24.1 | M12 |
| P01024 | M2 | SL_SM.C16.0 | M12 |
| P01031 | M2 | SL_SM.C16.1 | M12 |
| P01042 | M2 | SL_SM.C18.0 | M12 |
| P02748 | M2 | SL_SM.C18.1 | M12 |
| P02749 | M2 | SL_SM.C24.0 | M12 |
| P02760 | M2 | SL_SM.C24.1 | M12 |
| P02763 | M2 | SL_SM.C26.1 | M12 |
| P02765 | M2 | GP_PC.aa.C42.0 | M13 |
| P02774 | M2 | GP_PC.aa.C42.1 | M13 |
| P02787 | M2 | GP_PC.ae.C42.4 | M13 |
| P04003 | M2 | GP_PC.ae.C42.5 | M13 |
| P04004 | M2 | GP_PC.ae.C44.4 | M13 |
| P04217 | M2 | GP_PC.ae.C44.5 | M13 |
| P04275 | M2 | GP_PC.ae.C44.6 | M13 |
| P05090 | M2 | GP_PC.aa.C32.1 | M14 |
| P06681 | M2 | GP_PC.aa.C34.1 | M14 |
| P07225 | M2 | GP_PC.aa.C36.1 | M14 |
| P07357 | M2 | GP_PC.ae.C34.0 | M14 |
| P07358 | M2 | GP_PC.ae.C34.1 | M14 |
| P08603 | M2 | GP_PC.ae.C36.0 | M14 |
| P10643 | M2 | GP_PC.ae.C36.1 | M14 |
| P10909 | M2 | GP_PC.ae.C36.2 | M14 |
| P13671 | M2 | A0A024R6I7 | M15 |
| P14151 | M2 | A0A087WTK0 | M15 |
| P18065 | M2 | A0A087X0M8 | M15 |
| P19320 | M2 | A0A0A0MRJ7 | M15 |
| P19652 | M2 | A0A0A0MSA9 | M15 |
| P20851 | M2 | A0A0A0MSV6 | M15 |
| P22692 | M2 | C9JXF9 | M15 |
| P23142 | M2 | D6RF35 | M15 |
| P25311 | M2 | E7END6 | M15 |
| P27918 | M2 | H0Y2X5 | M15 |
| V9GYE7 | M2 | I3L0A1 | M15 |
| P41222 | M2 | I3L145 | M15 |
| P43652 | M2 | O14791 | M15 |
| P48740 | M2 | O75636 | M15 |
| P55056 | M2 | P01008 | M15 |
| Q03591 | M2 | P01011 | M15 |
| Q04756 | M2 | P01019 | M15 |
| Q12805 | M2 | P01023 | M15 |
| Q14520 | M2 | P01034 | M15 |
| Q16610 | M2 | P02647 | M15 |
| Q6UXB8 | M2 | P02649 | M15 |
| GP_lysoPC.a.C28.0 | M3 | V9GYM3 | M15 |
| GP_PC.aa.C36.0 | M3 | P02743 | M15 |
| GP_PC.aa.C38.0 | M3 | P02745 | M15 |
| GP_PC.aa.C40.1 | M3 | P02747 | M15 |
| GP_PC.aa.C40.2 | M3 | P02750 | M15 |
| GP_PC.aa.C40.3 | M3 | P02751 | M15 |
| GP_PC.aa.C42.2 | M3 | Q5VY30 | M15 |
| GP_PC.aa.C42.4 | M3 | P02766 | M15 |
| GP_PC.aa.C42.6 | M3 | P02775 | M15 |
| GP_PC.ae.C38.0 | M3 | P02776 | M15 |
| GP_PC.ae.C38.2 | M3 | P02790 | M15 |
| GP_PC.ae.C38.3 | M3 | P04075 | M15 |
| GP_PC.ae.C40.1 | M3 | P04114 | M15 |
| GP_PC.ae.C40.2 | M3 | P04196 | M15 |
| GP_PC.ae.C40.3 | M3 | P04264 | M15 |
| GP_PC.ae.C42.1 | M3 | P05155 | M15 |
| GP_PC.ae.C42.2 | M3 | P05362 | M15 |
| GP_PC.ae.C42.3 | M3 | P05452 | M15 |
| GP_PC.ae.C44.3 | M3 | P05543 | M15 |
| GP_PC.aa.C32.2 | M4 | P05546 | M15 |
| GP_PC.aa.C32.3 | M4 | P06396 | M15 |
| GP_PC.aa.C34.2 | M4 | P06727 | M15 |
| GP_PC.aa.C34.3 | M4 | P07360 | M15 |
| GP_PC.aa.C34.4 | M4 | P07996 | M15 |
| GP_PC.aa.C36.2 | M4 | P08185 | M15 |
| SL_SM.C20.2 | M4 | P08571 | M15 |
| AA_Ala | M5 | P08697 | M15 |
| AA_Gly | M5 | P0C0L4 | M15 |
| AA_Ile | M5 | P0C0L5 | M15 |
| AA_Leu | M5 | P10451 | M15 |
| AA_Lys | M5 | P13796 | M15 |
| AA_Met | M5 | P15169 | M15 |
| AA_Orn | M5 | P19823 | M15 |
| AA_Phe | M5 | P19827 | M15 |
| AA_Pro | M5 | P21333 | M15 |
| AA_Thr | M5 | P22792 | M15 |
| AA_Trp | M5 | P27169 | M15 |
| AA_Tyr | M5 | P29622 | M15 |
| AA_Val | M5 | P33151 | M15 |
| BA_Kynurenine | M5 | P36955 | M15 |
| AC_C5.DC..C6.OH. | M6 | P43251 | M15 |
| AC_C9 | M6 | P51884 | M15 |
| AC_C14.1 | M6 | P60174 | M15 |
| AC_C16.1 | M6 | P60709 | M15 |
| AC_C16.2 | M6 | P61769 | M15 |
| AC_C18 | M6 | P62328 | M15 |
| AC_C18.1 | M6 | P62937 | M15 |
| AC_C18.2 | M6 | Q06033 | M15 |
| A0A024R0T9 | M7 | Q08380 | M15 |
| A0A096LPE2 | M7 | Q13201 | M15 |
| B0YIW2 | M7 | Q14624 | M15 |
| E9PFZ2 | M7 | Q15582 | M15 |
| O00391 | M7 | Q96PD5 | M15 |
| P00738 | M7 | Q9NZP8 | M15 |
| P02671 | M7 | A0A0A0MRZ8 | M16 |
| P02675 | M7 | A0A0A0MS08 | M16 |
| P02679 | M7 | A0A0B4J231 | M16 |
| P02741 | M7 | S4R460 | M16 |
| P04180 | M7 | A0A286YES1 | M16 |
| P05109 | M7 | A0A286YFY1 | M16 |
| P06702 | M7 | A0A286YFY4 | M16 |
| P0DJI8 | M7 | A0A2Q2TTZ9 | M16 |
| P0DJI9 | M7 | O43866 | M16 |
| AA_Asn | M7 | P01591 | M16 |
| AA_Asp | M7 | P01619 | M16 |
| AA_Cit | M7 | P01706 | M16 |
| AA_Gln | M7 | P01780 | M16 |
| AA_His | M7 | P01834 | M16 |
| BA_Ac.Orn | M7 | P01871 | M16 |
| BA_alpha.AAA | M7 | P0DOY3 | M16 |
| BA_Creatinine | M7 | P80748 | M16 |
| BA_Met.SO | M7 |  |  |
| BA_Putrescine | M7 |  |  |
| BA_Serotonin | M7 |  |  |
| BA_Spermidine | M7 |  |  |
| BA_Spermine | M7 |  |  |
| BA_t4.OH.Pro | M7 |  |  |
| BA_Taurine | M7 |  |  |
| AC_C0 | M7 |  |  |
| AC_C2 | M7 |  |  |
| AC_C3 | M7 |  |  |
| AC_C3.DC..C4.OH. | M7 |  |  |
| AC_C4 | M7 |  |  |
| AC_C5 | M7 |  |  |
| AC_C10 | M7 |  |  |
| AC_C12 | M7 |  |  |
| Hexose | M7 |  |  |
| OA_Lactic.acid | M7 |  |  |
| OA_beta.Hydroxybutyric.acid | M7 |  |  |
| OA_alpha.Ketoglutaric.acid | M7 |  |  |
| OA_Citric.acid | M7 |  |  |
| OA_Butyric.acid | M7 |  |  |
| OA_Propionic.acid | M7 |  |  |

|  |  |
| --- | --- |
| OA_HPHPA | M7 |
| OA_Succinic.acid | M7 |
| OA_Fumaric.acid | M7 |
| OA_Pyruvic.acid | M7 |
| OA_Isobutyric.acid | M7 |
| OA_Hippuric.acid | M7 |
| OA_Methylmalonic.acid | M7 |
| OA_Homovanillic.acid | M7 |
| OA_Indole.acetic.acid | M7 |
| OA_Uric.acid | M7 |
| OA_5.Hydroxyindole.acetic.acid | M7 |
| OA_p.Hydroxyphenylacetic.acid | M7 |

